# A realist review of medication optimisation of community dwelling service users with serious mental illness

**DOI:** 10.1101/2023.08.02.23293542

**Authors:** Jo Howe, Maura MacPhee, Claire Duddy, Hafsah Habib, Geoff Wong, Simon Jacklin, Katherine Allen, Sheri Oduola, Rachel Upthegrove, Max Carlish, Emma Patterson, Ian Maidment

**Author notes:** Joint first authors (Jo Howe, Maura MacPhee).

## Abstract

**Background:** Severe mental illness (SMI) incorporates schizophrenia, bipolar disorder, non-organic psychosis, personality disorder or any other severe and enduring mental health illness. Medication, particularly anti-psychotics and mood stabilisers are the main treatment options. Medication optimisation is a hallmark of medication safety, characterized by the use of collaborative, person-centred approaches. There is very little published research describing medication optimisation with people living with SMI.

**Objective:** Published literature and two stakeholder groups were employed to answer: What works for whom and in what circumstances to optimise medication use with people living with SMI in the community?

**Methods:** A five-stage realist review was co-conducted with a lived experience group of individuals living with SMI and a practitioner group caring for individuals with SMI. An initial programme theory was developed. A formal literature search was conducted across eight bibliographic databases, and literature were screened for relevance to programme theory refinement. In total 60 papers contributed to the review. 42 papers were from the original database search with 18 papers identified from additional database searches and citation searches conducted based on stakeholder recommendations.

**Results:** Our programme theory represents a continuum from a service user’s initial diagnosis of SMI to therapeutic alliance development with practitioners, followed by mutual exchange of information, shared decision-making and medication optimisation. Accompanying the programme theory are 11 context-mechanism-outcome configurations that propose evidence-informed contextual factors and mechanisms that either facilitate or impede medication optimisation. Two mid-range theories highlighted in this review are supported decision-making and trust formation.

**Conclusions:** Supported decision-making and trust are foundational to overcoming stigma and establishing ‘safety’ and comfort between service users and practitioners. Avenues for future research include the influence of stigma and equity across cultural and ethnic groups with individuals with SMI; and use of trained supports, such as peer support workers.

**What is already known on this topic:** Medication optimisation is challenging for both people living with SMI and their prescribing clinicians; medication non-adherence is common.

**What this study adds:** Effective medication optimisation requires a person-centred approach embedded throughout a service user’s journey from initial diagnosis to effective medication co-management with practitioners.

**How this study might affect research, practice or policy:** Research is needed in multiple aspects of medication optimisation, including transition from acute care to community, the role of trained peer support workers, and practitioner awareness of unique needs for individuals from ethnic and cultural minority groups.

## INTRODUCTION

### Medication Optimisation

Severe mental illness (SMI) is a significant global healthcare burden with rates increasing throughout the world.^1^ The term SMI includes diagnoses of schizophrenia, non-organic psychosis, bipolar disorder, personality disorder and any other severe and enduring mental illness.^2^ Medications are a key treatment for SMI, but medication side-effects can contribute to chronic physical illness (e.g., diabetes, cardiovascular disease), a diminished quality of life and a decreased lifespan^3 4^. Complex medication regimens are often used to treat SMI; dosing can be a delicate balance between over or under-prescribing, based on individual service users’ (SUs) unique needs.^5^

Given the complex nature of SMI medication management and the need to consider issues such as risk of relapse, serious side effect profiles, and potential drug-drug interactions, medication safety is of paramount importance to SUs and practitioners^6–8^. Since 2008, the global Institute for Healthcare Improvement (IHI) and its country affiliates, such as the UK’s National Health Foundation, have advocated for inclusion of service users (SUs) and person-centred care approaches when identifying best practices and strategies pertaining to patient safety and quality of care delivery.

The original Triple Aim IHI framework consisted of three pillars for advancing quality and safety: enhanced population health, positive SU experiences, and cost-effectiveness.^9^ The original framework has expanded to Quadruple Aim, including staff experience.^9^ These IHI frameworks highlight how the SU voice is an integral component of all healthcare quality and safety initiatives. Medication optimisation, a hallmark of medication safety, is defined as “a person-centred approach to safe and effective medicines use”.^10^ Effective medication optimisation involves the perspectives of SUs with lived experience of taking medications^11^ and multi-disciplinary care delivery for SMI is more effective when SUs play a central role in medication decision-making.^12^

Failure to optimise medication is often attributed to SU non-adherence, practitioner under or over-prescribing or over-treatment, including polypharmacy.^2 12–14^ Management of SMI is particularly challenging with reported non-adherence rates as high as 50%.^15 16^ Non-adherence^17^, and over-prescribing occur more frequently in ethnic minority communities, as do physical illnesses, such as diabetes and cardiovascular disease.^10 18^ In general, the lowest possible medication dose is recommended to control SMI symptoms^19^, however higher doses are often prescribed by practitioners concerned about relapse^20^. Poorly managed SMI increases relapse rates, hospitalisation and is associated with unemployment, homelessness, disrupted education, substance misuse, physical health problems, self-harm and excess mortality^2 13 14 21^. A systematic review and meta-analysis of studies from Asia, Europe and North America found that non-adherence within the SMI population is the strongest predictor of relapse.^22^

### Shared Decision-making

Person-centred approaches, such as shared decision-making (SDM) between key practitioner groups (e.g., pharmacy, medicine, nursing), SUs with SMI, and family carers, are associated with increased SU medication adherence and improved practitioner prescribing practices. ^7 13 15 16^ There is, however, limited research on what needs to happen, how and when in the SU-practitioner relationship to promote person-centred SDM, and ultimately, medication optimisation for SUs with SMI. ^4 13 23^ Assumptions are often made about intervention effectiveness only from practitioner’s viewpoints.^24^ The implementation of SDM can be hindered by practitioners’ beliefs about SDM. A Netherlands based study compared practitioner reports of SDM use with direct observations of their SU interactions.^25^ Practitioners reported using SDM as their usual decision-making style, but in observations, there was low engagement with SUs. The authors described practitioners as “unconsciously incompetent in SDM”. Therefore, developing knowledge on how to implement effective patient centred approaches that promote medication optimisation is needed."

### Present Research

We conducted a realist review on medication optimisation with community dwelling SU’s living with SMI. We focussed on community dwelling as most SMI service users live in the community; where there are greater opportunities for them to exercise control over their medication regime (e.g., by omitting doses, or via non-adherence).

We synthesised data from academic literature and drew on perspssectives of community-based SUs stakeholders with lived experience of SMI, informal (family) carers and mental health practitioners caring for SU’s with SMI. A realist review can uncover important contextual factors affecting outcomes.^26^ We constructed a programme theory comprised of a series of testable hypotheses, known as context-mechanism-outcome configurations (CMOCs), to explain a potential SU-practitioner journey from initial diagnosis to trusting therapeutic alliance, shared decision-making and medication optimisation.

Realist reviews have become increasingly popular within the quality and safety literature to explain how and why interventions work. Realist reviews have been used to investigate junior doctors’ anti-microbial prescribing^27^; safety-netting practices in primary care^28^; and medication management for community-dwelling seniors on complex medication regimens.^29^ Realist reviews address research questions about what works, for whom, under what circumstances and how, and are a valuable methodological alternative or complement to other forms of evidence synthesis, such as systematic review.^26 30^

### Research objectives

The overall aim for this realist review was to use published literature, alongside lived experience and practitioner stakeholder groups to understand: What works for whom and in what circumstances to optimise medication for community dwelling SUs with SMI.

## METHODS

We conducted a five-stage realist review. Our review protocol was published^31^ and registered with PROSPERO (CRD42021280980). We used academic literature as well as feedback and advice from our stakeholder groups (LEG [Lived Experience Group], and PG [Practitioner Group]) to refine our programme theory, and create a series of testable CMOCs.^32^ The LEG comprised 6 lived experience stakeholders from Birmingham and Solihull Mental Health Foundation Trust (BSMHFT) Lived Experience Advisory Research (LEAR) Group and 2 additional individuals with lived experience from outside LEAR (who were recruited to facilitate discussions). The PG comprised healthcare practitioners from the United Kingdom caring for individuals with SMI. The practitioners were recruited from personal networks and via social media advertisements. Our CMOCs describe specific contexts associated with important outcomes related to medication optimisation, such as therapeutic alliance formation, and to articulate the mechanisms that trigger these outcomes. In this project, we have defined the following:

**Context:** Adults living with SMI on medication

**Intervention:** any intervention to optimise medication usage; people living with SMI, family carers’ and practitioners’ experiences of managing and using medication.

**Mechanisms:** hidden, psychological processes that link specific contexts to intended outcomes.

**Outcomes:** quality of life, adherence, adverse events, disease symptoms, economic.

### Stage 1: Objectives, initial programme theory

#### Objectives

To conduct a realist review using published literature, alongside lived experience and practitioner stakeholder input to understand: What works for whom and in what circumstances to optimise medication for community dwelling SUs with SMI.

#### Development of initial programme theory

We developed an initial programme theory (IPT), a testable explanation of how and why medication optimisation, is supposed to work for people living with SMI. This IPT was informed by an initial informal literature search, stakeholder engagement and subject matter experts known to the project team.

### Stage 2: Literature search

A formal literature search was conducted in January 2022 by CD across eight bibliographic databases (MEDLINE, Embase, PsycINFO, CINAHL, Cochrane Library, Scopus, Web of Science Core Citation Indexes [SCIE, SSCI, SHCI, ESCI, CPCI, BKCI] and Sociological Abstracts). Our searches combined free text and subject heading terms for SMI, with terms describing medication or medication optimisation, and a comprehensive list of terms reflecting our project focus on SDM and SU-practitioner relationships. The LEG and PG helped to identify key concepts used in our search strategy. Our original protocol indicated that we would run searches in Google Scholar, but this was deemed unnecessary following screening, in light of the volume of literature already retrieved. In response to PG and LEG feedback, additional targeted searches were conducted in June 2022 to identify material relating to internet use, peer support and tapering medication.

Full details of our search strategies are available in supplementary File 1.

### Stage 3: Screening and inclusion

Inclusion criteria focussed on community dwelling adults (18+) living with SMI and taking antipsychotic medication. Studies limited to inpatient settings or focused on diagnoses outside of SMI were excluded. Full details of the inclusion/exclusion criteria can be found in Table 1.

**Table 1:** Review inclusion and exclusion criteria.

| **Inclusion** | **Exclusion** |
| --- | --- |
| Community dwelling adults living with SMI, using medication.<br>Their family or carers.<br>Practitioners involved in their care | Inpatient settings only |
| Interventions to optimise medication usage<br>Or<br>Experience of medication management and use | Shared decision-making tools |
| All study designs | No focus on shared decision-making process |
| All countries | Not SMI |
| English language | Eating Disorders |

#### Screening

The results of the main search were screened by title and abstract by JH using RAYYAN (a web-based tool designed to assist with screening of title and abstract). A random 10% sample was screened in duplicate by MM. Uncertainties were resolved via discussions with JH, and MM. Full text screening was initially completed in EndNote X9 by JH on documents published from 2014 onwards. The decision to focus on this timepoint was based on a significant increase in documents with SDM content during this time period. All documents within Endnote were assigned a star rating of one to five by JH, based on a global judgement of each documents’ likely relevance, richness and rigour. One-star documents were deemed irrelevant and rejected. Two-star documents were deemed “unsure”, these were subsequently discussed with MM and reallocated. Three-star documents were deemed irrelevant for programme theory development but potentially for background. Four-star documents were deemed relevant to CMOC development and programme theory refinement. Five-star documents were deemed the most conceptually rich and most relevant to CMOC development and programme theory refinement.

Pre 2014 documents and documents obtained via citation searching and personal networks were purposively screened and analysed by CD, JH, MM and HH but were not categorised with a star rating as they were chosen due to perceived high relevance, richness and rigour.

### Stage 4: Data extraction, analysis

#### Data extraction

Document characteristics were extracted to an Excel spreadsheet by HH (supplementary file 2). A coding framework was iteratively and inductively developed and tested by MM, JH and HH to organise relevant data (supplemental file 3).

#### Data analysis

Coding of post-2014 five-star full text documents was completed in NVivo by MM and HH with a 10% check in duplicate by JH. Extracts of data were coded to nodes (termed parent nodes in NVivo) reflecting conceptual buckets e.g., SDM, independent decision-making, therapeutic alliance, coercion etc. Extracts of data were coded against sub-nodes (called child nodes in NVivo) and multiple nodes if appropriate.

### Stage 5: Data synthesis, CMOC development and programme theory refinement

Once all five-star papers had been coded and discussed with the PG and LEG, and due to the time limited nature of the review, a pragmatic decision was made to narrow the focus of the review on decision-making and therapeutic alliance. Coded data relating to SDM, and therapeutic alliance were initially extracted and imported in Microsoft Word by MM and HH and CMOCs were initially developed through ongoing discussions with JH. These CMOCs were iteratively refined by checking remaining data from NVivo nodes and extracting relevant examples. Further refinement of the programme theory and CMOCs occurred using data from relevant four-star papers, pre-2014 papers, papers from additional searches and discussions with the project team, LEG and PG. The finalised set of CMOCs and a refined programme theory were discussed and validated with the PG, the LEG, and the wider project team. The refined CMOCs, supporting evidence, and document origin (e.g., post-2014, via citation search or personal networks) can be found in supplemental file 4.

## FINDINGS

Our main search identified 1118 unique results. After title and abstract screening, 144 documents published from 2014 onwards, were screened in full text. 29 papers were assigned a five-star rating and coded in NVivo. 33 papers were assigned a four-star rating. Following the decision to narrow the focus, of these 62 papers, 27 were rejected leaving a total of 35 four and five-star papers in the review. 18 papers were identified via additional searches, citation searches and personal contacts. 7 of the pre-2014 papers were deemed relevant taking the total number of papers contributing to the review to 60 (35 + 18 + 7). Our searching and screening processes are summarised in Figure 1 below.

**Figure 1:**
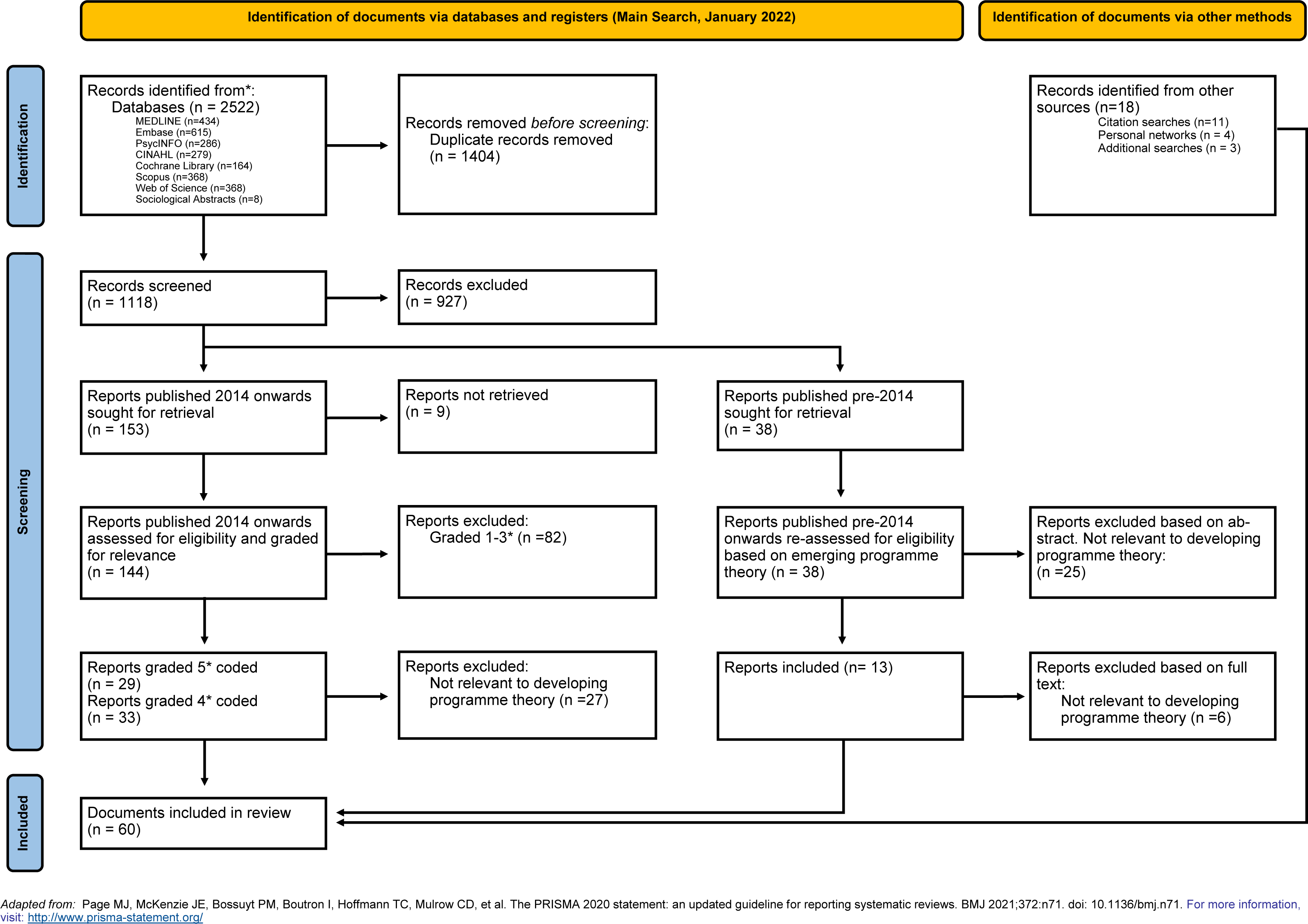
PRISMA 2020 flow diagram for new systematic reviews which included searches of databases, registers and other sources.

Table 2 includes our refined programme theory and 11 CMOCs underpinning the theory. Our refined programme theory describes a journey of medication optimisation for individuals with SMI that begins with initial diagnosis and culminates in a therapeutic alliance characterised by underlying trust, mutual information exchange and shared decision-making. There are potential barriers and facilitators along the way, represented by positive and negative CMOCs. The journey includes practitioners, SUs with their family and social network, and other information sources (e.g., Internet, peer support workers).

**Table 2.** Refined Programme Theory and underpinning Context-Mechanism-Outcome Configurations.

| **Refined Programme Theory** |  |
| --- | --- |
| *“When service users (SU) are first diagnosed with serious mental illness (SMI), a diagnosis which is frightening to them, they seek out information about their illness.*<br>*SUs on medications want practitioner support and practitioner advice they can understand and apply to their current and ongoing needs.*<br>*SUs may seek out individuals with lived experience to validate the experiences they are having, and to learn how others effectively manage living with SMI. As SUs gather information from diverse sources (practitioners, social supports, Internet), they are constantly weighing up the pros and cons of medication decisions.*<br>*It is important to SUs to forge positive working relationships with practitioners who will listen to them, respectfully consider their needs, and support their medication decisions whenever possible. SUs are regularly facing lifestyle challenges, some with high stakes, such as pregnancy or serious health side effects. If and when SUs have established therapeutic relationships with practitioners who have their best interests at heart and are competent in their field of expertise, SUs are more apt to seek them out for shared information exchange and decision-making.*<br>*Regardless of the strength of the SU-practitioner relationship, in high stakes situations, trust is fragile; trust is based on ongoing evidence of practitioners’ motivations to support them. Similarly, SUs need ongoing and non-judgmental support from family members and their social network, including peer support workers.”* |  |
| **Context-Mechanism-Outcome-Configuration** | **Evidence Sources:** |
| **CMOC 1 First Contact**<br>When an individual with SMI is first diagnosed, is medicated and has coercive, dehumanizing* experiences with practitioners (C), this often derails the development of trusting therapeutic alliances (O) because of feelings of powerlessness (M) and stigmatization (M).<br>*Our lived experience group (LEG) asked us to include “dehumanizing,” based on their initial experiences with SMI diagnosis and management. | 33-42 |
| **CMOC 2 Relief**<br>When an individual with SMI is first diagnosed and is medicated, validation and normalization of their condition by a respectful, supportive practitioner (C) results in increased relief, hope and optimism (O) due to decreased stigmatization of living with SMI (M) and increased reassurance (M) that they have a treatable condition. | 36 37 40 43-45 |
| **CMOC 3 Dismissal**<br>When an individual with SMI on medications realizes practitioners are withholding medication information, and/or excluding, ignoring or dismissing them from medication decisions (C), they are apt to withdraw from the practitioner relationship and make their own medication decisions (O), due to mistrust (M) in the practitioners’ interest in them and their need for more control (M) over decisions affecting their lives. | 33 35 37 38 40 41 43-56 |
| **CMOC 4 Being heard**<br>From the start of their relationship onwards, when an individual with SMI on medications is actively engaged by a respectful, supportive practitioner who takes an interest in them and their issues and concerns about their illness, medication and side effects (C), they are more apt to forge a therapeutic alliance with their practitioner (O), because they feel heard and listened to (M) and they trust (M) in the practitioner's motivations to help them better manage their medications and illness. | 35 37 40 43 45 49 57-67 |
| **CMOC 5 Practitioner information exchange**<br>From the start of the therapeutic relationship onwards, when an individual with SMI feels comfortable accessing their practitioner for honest, easy-to-understand and personalized information about their medications (C), they are apt to use the information to prepare for and to cope better with medications and side effects (O), due to development of mutual trust (M) and respect (M) in each other and in the information being exchanged. | 33 38 41-43 45 46 48 49 52-54 57 59 68-72 |
| **CMOC 6 Seeking more information**<br>Whenever an individual with SMI on medications desires additional information about their illness, medications and potential side effects (C), they will often seek out accessible, easy-to-understand information from a variety of non-practitioner sources (e.g., peers, Internet) they perceive to be trustworthy and credible (O), due to need for increased knowledge (M) increased reassurance (M) and greater control (M) with respect to medication and life decisions. | 38 40 44 46 48 51 52 57 70 73 |
| **CMOC 7 Confiding and negotiating in a safe way**<br>When an individual with SMI on medications has continuity over time in a respectful, trusting therapeutic alliance with practitioners who openly discuss and make collaborative medication decisions with them, even when there are disagreements (C), they are more apt to confide in and to negotiate with their practitioners about their medication issues and management plans (O), due to a sense of safety with their practitioners (M), and increased belief (M) in themselves to manage their lives. | 33-36 40 42-50 57 58 64 66 67 70-72 74-83 |
| **CMOC 8 Perceived Risks**<br>When individuals with SMI desire to taper, change or discontinue their medication regimen (C), their clinicians may resist sharing information with them and may not support them (O) because they judge that doing so may put themselves, the patient and others at risk (M) if adverse outcomes occur (e.g., harm to self or others). | 37 41 47 49 52-54 71 74 84 85 |
| **CMOC 9 Family and Social Supports**<br>When an individual with SMI has support from family and social network members who believe in them and want the best for them (C), they are apt to feel more confident in following through with prescribed medication plans (O) due to increased belief (M) in their capacity to handle ongoing challenges and a sense of safety (M) among people looking after their well-being. | 50 52 62 64 77 79 86-88 |
| **CMOC 10 Fear and Guilt** | 37 40 48 50 51 62 64 79 87 89 90 |
| When an individual with SMI is aware that their family members are fearful about the consequences from medication changes and want them to maintain medications as prescribed (C), they may continue on the medications against their will or secretly discontinue/change their medications (O) to avoid conflict (M) and/or withdrawal of their family's support (M) for them. |  |
| **CMOC 11 Peer Support Workers (PSWs)**<br>When individuals with SMI have access to peer support workers with shared lived experiences who talk with them about SMI and life skills management, including medications and side effects (C), they are apt to experience a positive impact on their mental, physical and social-emotional health (O) because they feel validated (M) less stigmatized (M) and reassured (M) that they can have a productive, fulfilling lives with SMI. | 35 52 58 75 91 92 |

In Table 1, CMOCs 1 (First contact) and 2 (Relief) are associated with initial diagnosis. The literature highlights the importance of positive first encounters with healthcare services. Negative, coercive experiences can derail practitioner-SU trust formation; while positive experiences can decrease internal stigma and reassure SUs that their condition is treatable.

CMOC 3 (Dismissal) depicts how dismissal and devaluing by practitioners of SUs impedes the establishment of trust, which is a foundational component to therapeutic relationships.

CMOC 4 (Being heard) illustrates how development of the therapeutic alliance is dependent on respectful, supportive practitioners willing to listen to and seriously consider SU’s needs and concerns.

CMOCs 5 (Practitioner information exchange) and 6 (Seeking more information) represent SUs’ desire for credible, trustworthy information about diagnosis and medication that is personalised to their living with SMI and the role of medication in treating their illness, including possible side effects. Regardless of information obtained from practitioners, SUs typically seek out additional information to obtain new knowledge, a greater sense of reassurance, and more control over medication and life decisions.

As described in CMOC 7 (Confiding and negotiating in a safe way), a hallmark of strong and effective therapeutic alliances is the ability of practitioners to support SUs, even if they disagree with their medication decisions. SUs feel safe in this type of alliance and are more apt to collaboratively plan their care with a trusted practitioner.

In contrast, CMOC 8 (Perceived risks), illustrates how some practitioners have difficulty supporting SUs’ wishes due to risk about potential adverse outcomes, such as relapse and its consequences.

CMOCs 9 (Family and social supports), 10 (Fear and guilt) and 11 (Peer supports) are related to non-practitioner sources of support for individuals with SMI. The family can be a safety net and positive support for SUs (CMOC 9), or the family can be fearful of making medication changes (CMOC 10), resulting in negative SU emotions, such as fear and guilt. In these situations, SUs may feel pressured to conform to family wishes to avoid conflict and ensure that family support is not withdrawn. Peer support workers (CMOC 11) are a promising source of support to SUs, because they validate SUs feelings, given their lived experience with SMI. However, research on peer support worker roles in medication optimisation was lacking.

## DISCUSSION

The patient safety literature has demonstrated that higher levels of safety are achievable by ensuring the voice of SUs and other stakeholders are part of quality improvement efforts.^93^ Medication optimisation is an important component of patient safety, especially for SUs with SMI where medication is the key treatment strategy. Our programme theory and CMOCs highlight how person-centred care approaches such as providing relevant, useful information and support through practitioners and others can lead to safe medication use (i.e. medication optimisation). The CMOCs outlined above provide testable, causal explanations for outcomes, detailing by whom, when and how these happen.

### Comparisons with existing literature and theory

A valuable aspect of the realist approach is the potential to use formal or substantive theories to further explain and buttress inferences about underlying mechanisms or drivers for individuals’ actions. Based on our reading of the included documents and recommendations from our stakeholder groups, we discuss below two theories of particular importance to therapeutic alliance development, which is a necessary condition for effective medication optimisation.

### Supported decision-making theory

Supported decision-making (SUDM) theory emphasises practitioner’s roles in assisting SUs with decision-making based on their needs and preferences. SUDM theory has legal and ethical associations with the 2006 United Nations Convention of Individuals’ Rights and Disabilities, which stipulated that no person should be appointed as a decision-maker for an individual who has the capacity to make their own decisions with appropriate supports.^94^ SUDM encompasses person-centred planning, advocacy, communications, interpretive supports and representational supports (e.g., peer support workers, family and social networks) and the central question practitioners should ask themselves is: “What supports are needed to ensure this person can best exercise their rights?”^94^

Although research into supported decision-making and SMI is rare, qualitative researchers have found that the timing and types of practitioner supports made a difference to individuals with SMI, particularly with respect to confidence and self-control.^95^ ^96^ Researchers from one study created different thematic labels for SUs: SUs with capacity to make their own cogent decisions were “inward experts;” while SUs during periods of acute unwellness were “outward entrustors,” entrusting practitioners to guide their care management. SUs’ variable needs for SUDM required different practitioner roles, such as practitioners as facilitators (e.g., sharing information openly and honestly) and as collaborators (e.g., promoting shared decision-making)^95^

In a recent systematic review of SUDM for SUs with SMI in clinical practice settings,^97^ a limited number of papers included in the review examined stakeholders’ perspectives of SUDM. Stakeholders, including SUs, family members and practitioners, all agreed to the importance of SUDM. Practitioner misconceptions about differences between SUs’ rights and preferences, however, were barriers to implementation success. If SUDM is a necessary condition for SU-practitioner SDM and medication optimisation, practitioners will need to understand SUs’ legal rights, and to engage in roles (e.g., facilitator, collaborator) that promote SUs’ decision-making autonomy^95^

### Trust formation theory

Our findings make clear that, for SUs with SMI, ongoing alliance-building and confidence in their capacity to share in decisions and manage their lives depends on trust: trust in practitioners and trust/belief in themselves. Trust formation theory defines trust as “a psychological state comprising the intention to accept vulnerability based upon positive expectations of the intentions or behaviour of another”.^98^ Our programme theory proposes that trust between SUs and practitioners evolves with the development of the therapeutic alliance. An exploratory study of the role of trust in medication management within mental health services ^99^ supported our realist review findings that practitioners’ reluctance to share useful information in an open and honest way can create mistrust and worsen medication adherence. Ultimately, mistrust obstructs collaborative medication management.^99 100^

The wider literature provides some evidence of how mutual trust formation enables engagement, disclosure and collaboration in mental health care. Corroborating our findings, a qualitative study from the UK found that practitioners’ open communications and therapeutic listening promoted and sustained the development of mutual trust over time.^101^ More recent literature suggests that the development of trusting therapeutic alliance is enhanced by practitioners’ awareness and respect for SU preferences, such as types of treatment options (e.g., medications, psychotherapy), and influenced by the characteristics of practitioners they work with (e.g., professional background, gender, age and ethnicity).^102^ Even when all preferences cannot be accommodated, eliciting, discussing and acknowledging SU preferences is associated with stronger alliances.^102^

### The relationship between SUDM, trust and information

CMOCs (4,5,7) associate SDM with active engagement of SUs-practitioners in open, transparent discussions and collaborative treatment planning based on mutual trust. CMOC 8, however, addresses the negative emotions and risks associated with the SU-practitioner relationship. The literature in our review focused predominantly on practitioner risks, such as concern for SU medication non-adherence and adverse SU outcomes. A recent review of qualitative studies^103^ discussed risk-taking from the perspectives of SUs and practitioners. With shared risk-taking, both parties jointly reflect and address inherent risks to any decision, particularly from a safety perspective. Some evidence suggests that mutually identifying and preventing or mitigating risks can actually strengthen the alliance and deepen trust.^104^ In the UK, the NHS recommends that risk assessment and management should be explained to SUs with SMI as soon as possible as part of SDM.^105^

CMOCs (6,9,11) pertain to non-practitioner sources of information and support, including the Internet and social media, family and friends, and peer supports with similar lived experience, although we identified very limited academic literature on how peer support workers can be used to optimise medication management.^35 52 58 75 91 92^ Questions exist with respect to peer support workers’ capacity to give accurate medication advice.^106^

### Future research directions

As mentioned above, future research needs to address how non-practitioner sources of information and supports, (e.g., peer support workers, families and friends, internet and social media). Our LEG and PG stakeholders both endorsed the importance of peer support workers and their potential roles in medication optimisation.

An area of burgeoning interest is online decision support tools to improve information sharing and communication between SUs and practitioners. A recent systematic review found mixed evidence for the effectiveness of decision support tools with SUs with SMI.^107^ This review included tools to assist with prioritising treatment preferences, crisis planning and advanced directives. More conclusive research is needed to evaluate the efficacy of online support tools, especially how they help or hinder therapeutic alliance development, SDM and medication optimisation for SU’s with SMI.

Our review has highlighted an important evidence gap relating to equity, diversity and inclusivity (EDI) for SUs with SMI from racial and ethnic minority groups, seniors and other vulnerable populations. These groups were rarely mentioned in our review’s included papers. When SUs with SMI are members of minority groups and/or vulnerable populations, stigma can be compounded.^23 108^ Ultimately, the success of SU-practitioner relationships depends on reducing mistrust among SUs who have been stigmatized by SMI and by race/ethnicity, while enhancing practitioners’ awareness and commitment to EDI. In England, the Race Equality Foundation is a national charity that tracks and reports racial inequality in public services (https://raceequalityfoundation.org.uk/). Researchers working with this charity identified persistent healthcare inequalities for English minority groups. “Traumatic, inappropriate and discriminatory experiences of services can have a detrimental impact on chances for recovery, particularly if the same risk factors of bereavement, family breakdown, incarceration, poverty and exposure to racism continue to be present.”^23^

Community-based care models may be a more cost-effective way of caring for complex and vulnerable patients. Only a limited number of new care models (e.g., team-based care, integrated care) have been well-described or evaluated for SUs with SMI.^109^ In one mapping review of integrated physical-mental care models for SUs with SMI, a number of concerns were identified, including practitioners’ negative biases and stigma towards SMI^110^. As our review illustrated, the initial context, particularly the presence of any negative biases towards SMI, can derail the development of a therapeutic alliance between SUs and practitioners.

### Strengths and limitations

We conducted our realist review within a one-year timeline. To be as efficient as possible, we focused on the largest body of relevant literature published from 2014 onwards. We returned to pre-2014 literature after developing our CMOCs from the more recent literature to identify earlier data relating to the CMOCs. Our review identified evidence gaps in relation to the relationship between race, ethnicity, vulnerable groups and medication optimisation in SMI and in the role of peer-to-peer support workers.

A strength of this review was the engagement with PG and LEG stakeholders throughout the review process. Their engagement supported our interpretations of data, ensured our findings reflected their real-world experiences and highlighted gaps in the literature. Realist reviews are an iterative process of developing theory and CMOCs, which may then be confirmed, refuted or refined by future research, including, for example, realist evaluation. This review’s programme theory and CMOCs produced testable hypotheses for future research with individuals with SMI and community-based practitioners who serve them.

### Conclusions

Medication optimisation is a medication safety ‘gold standard’ for SUs with SMI, given the physical and mental health sequelae associated with non-adherence and over-or under-prescribing. Medication optimisation is a person-centred approach that begins at time of initial diagnosis and ensures optimal information and supports are accessible to SUs, based on their needs and preferences. Practitioner actions need to promote the SU voice in all aspects of their recovery journey.

## Supporting information

Supplemental File 1, and will be used for the link to the file on the preprint slide

Supplemental File 2, and will be used for the link to the file on the preprint slide

Supplemental File 3, and will be used for the link to the file on the preprint slide

## Data Availability

All data produced in the present study are available upon reasonable request to the authors.

## Acknowledgements

We would like to thank our practitioner and lived experience stakeholder groups for their time and effort in helping us shape this research. Members of the lived experience group included but not limited to Max Carlish, Hameed Khan, Mustak Mirza, Barbara Norden, Janelle Peacock, Ian Tighe and Rosemary.

## Funding Disclaimer

This study/project is funded by the NIHR Programme Development Award (203683). The views expressed are those of the author(s) and not necessarily those of the NIHR or the Department of Health and Social Care.

## References

1. Jaeschke K, Hanna F, Ali S, et al. Global estimates of service coverage for severe mental disorders: findings from the WHO Mental Health Atlas 2017. Global Mental Health 2021;8:e27. doi: https://doi.org/10.1017/gmh.2021.19

2. NHS England. Improving physical healthcare for people living with severe mental illness (SMI) in primary care: guidance for CCGs 2018 [Available from: www.england.nhs.uk/wp-content/uploads/2018/02/improving-physical-health-care-for-smi-in-primary-care.pdf18.

3. Carolan A, Keating D, Strawbridge J, et al. Optimising prescribing for patients with severe mental illness: the need for criteria. Evidence-based Mental Health 2019;22(4):139–41. doi: http://dx.doi.org/10.1136/ebmental-2019-300099

4. National Institute for Health and Care Excellence. Psychosis and schizophrenia in adults: prevention and management 2014 [Available from: https://www.nice.org.uk/guidance/cg178.

5. Ostuzzi G, Vita G, Bertolini F, et al. Continuing, reducing, switching, or stopping antipsychotics in individuals with schizophrenia-spectrum disorders who are clinically stable: a systematic review and network meta-analysis. The Lancet Psychiatry 2022 doi: https://doi.org/10.1016/S2215-0366(22)00158-4

6. Davis KAS, Farooq S, Hayes JF, et al. Pharmacoepidemiology research: delivering evidence about drug safety and effectiveness in mental health. The Lancet Psychiatry 2020;7(4):363–70. doi: https://doi.org/10.1016/S2215-0366(19)30298-6

7. Maidment ID, Lelliott P, Paton C. Medication errors in mental healthcare: a systematic review. Quality and Safety in Health Care 2006;15(6):409–13. doi: http://dx.doi.org/10.1136/qshc.2006.018267

8. Roughead L, Procter N, Westaway K, et al. Medication safety in mental health. Sydney: Australian Commission on Safety and Quality in Health Care, 2017.

9. Obucina M, Harris N, Fitzgerald JA, et al. The application of triple aim framework in the context of primary healthcare: A systematic literature review. Health Policy 2018;122(8):900–07. doi: https://doi.org/10.1016/j.healthpol.2018.06.006

10. National Institute for Health and Care Excellence. Medicines optimisation: the safe and effective use of medicines to enable the best possible outcomes 2015 [Available from: https://www.nice.org.uk/guidance/ng5.

11. Lee JL, Dy SM, Gurses AP, et al. Towards a More Patient-Centered Approach to Medication Safety. Journal of Patient Experience 2018;5(2):83–87. doi: https://doi.org/10.1177/2374373517727532

12. Maidment I, Sud D, Chew-Graham C. Challenge of optimising medication in people with severe mental illness. BMJ Quality & Safety 2021;31(5):337–39. doi: https://doi.org/10.1136/bmjqs-2021-013847

13. Mental Health Taskforce. The five year forward view for mental health. 2016

14. Robotham D, Wykes T, Rose D, et al. Service user and carer priorities in a Biomedical Research Centre for mental health. Journal of Mental Health 2016;25(3):185–88. doi: https://doi.org/10.3109/09638237.2016.1167862

15. National Institute for health and Clinical Excellence. Medicines Adherence: Involving patiens in decisions about prescribed medicines and supporting adherence. 2009 [Available from: https://www.nice.org.uk/guidance/cg76.

16. Semahegn A, Torpey K, Manu A, et al. Psychotropic medication non-adherence and its associated factors among patients with major psychiatric disorders: a systematic review and meta-analysis. Systematic Reviews 2020;9(1):17. doi: https://doi.org/10.1186/s13643-020-1274-3

17. Opolka JL, Rascati KL, Brown CM, et al. Role of ethnicity in predicting antipsychotic medication adherence. Annals of Pharmacotherapy 2003;37(5):625–30. doi: https://doi.org/10.1345/aph.1C321

18. Sud D, Laughton E, McAskill R, et al. The role of pharmacy in the management of cardiometabolic risk, metabolic syndrome and related diseases in severe mental illness: a mixed-methods systematic literature review. Systematic Reviews 2021;10(1):92. doi: https://doi.org/10.1186/s13643-021-01586-9

19. Correll CU, Martin A, Patel C, et al. Systematic literature review of schizophrenia clinical practice guidelines on acute and maintenance management with antipsychotics. Schizophrenia 2022;8(1):5. doi: 10.1038/s41537-021-00192-x

20. Rodolico A, Siafis S, Bighelli I, et al. Antipsychotic dose reduction compared to dose continuation for people with schizophrenia. Cochrane Database of Systematic Reviews 2022(11) doi: https://doi.org/10.1002/14651858.CD014384.pub2

21. Weinmann S, Read J, Aderhold V. Influence of antipsychotics on mortality in schizophrenia: Systematic review. Schizophrenia Research 2009;113(1):1–11. doi: https://doi.org/10.1016/j.schres.2009.05.018

22. Loots E, Goossens E, Vanwesemael T, et al. Interventions to Improve Medication Adherence in Patients with Schizophrenia or Bipolar Disorders: A Systematic Review and Meta-Analysis. International Journal of Environmental Research and Public Health 2021;18(19):10213. doi: https://doi.org/doi:10.3390/ijerph181910213

23. Bignall T, Jeraj S, Helsby E, et al. Racial disparities in mental health: Literature and evidence review. London: Race Equality Foundation, 2019.

24. Haverfield MC, Tierney A, Schwartz R, et al. Can Patient–Provider Interpersonal Interventions Achieve the Quadruple Aim of Healthcare? A Systematic Review. Journal of General Internal Medicine 2020;35(7):2107–17. doi: https://doi.org/10.1007/s11606-019-05525-2

25. Ellen M. Driever, Anne MS, Paul LPB. Do consultants do what they say they do? Observational study of the extent to which clinicians involve their patients in the decision-making process. BMJ Open 2022;12(1):e056471. doi: https://doi.org/10.1136/bmjopen-2021-056471

26. Duddy C, Wong G. Grand rounds in methodology: when are realist reviews useful, and what does a ‘good’ realist review look like? BMJ Quality & Safety 2022;32(3):173–80. doi: https://doi.org/10.1136/bmjqs-2022-015236

27. Papoutsi C, Mattick K, Pearson M, et al. Social and professional influences on antimicrobial prescribing for doctors-in-training: a realist review. Journal of Antimicrobial Chemotherapy 2017;72(9):2418–30. doi: https://doi.org/10.1093/jac/dkx194

28. Friedemann Smith C, Lunn H, Wong G, et al. Optimising GPs’ communication of advice to facilitate patients’ self-care and prompt follow-up when the diagnosis is uncertain: a realist review of ‘safety-netting’ in primary care. BMJ Quality & Safety 2022;31(7):541–54. doi: https://doi.org/10.1136/bmjqs-2021-014529

29. Maidment I, Lawson S, Wong G, et al. Medication management in older people: the MEMORABLE realist synthesis. Health Services and Delivery Research, 2020:1–128.

30. Peytremann-Bridevaux I, MacPhee M. Moving Forward With Integrated Care: The Use of Realist Approaches to Understand What Works, How, for Whom and Under Which Circumstances. Public Health Reviews 2022;43 doi: https://doi.org/10.3389/phrs.2022.1605082

31. Maidment I, Wong G, Duddy C, et al. Medication optimisation in severe mental illness (MEDIATE): protocol for a realist review. BMJ Open 2022;12(1):e058524. doi: https://doi.org/10.1136/bmjopen-2021-058524

32. Wong G, Westhorp G, Manzano A, et al. RAMESES II reporting standards for realist evaluations. BMC Medicine 2016;14(1):96. doi: https://doi.org/10.1186/s12916-016-0643-1

33. Bolden GB, Angell B, Hepburn A. How clients solicit medication changes in psychiatry. Sociology of Health & Illness 2019;41(2):411–26. doi: https://doi.org/10.1111/1467-9566.12843

34. Carrick R, Mitchell A, Powell RA, et al. The quest for well_-_being: A qualitative study of the experience of taking antipsychotic medication. *Psychology and Psychotherapy: Theory*, Research and Practice 2004;77(1):19–33. doi: https://doi.org/10.1348/147608304322874236

35. Ehrlich C, Dannapfel P. Shared decision making: People with severe mental illness experiences of involvement in the care of their physical health. Mental Health and Prevention 2017;5:21–26. doi: https://doi.org/10.1016/j.mhp.2016.12.002

36. Maj M, van Os J, De Hert M, et al. The clinical characterization of the patient with primary psychosis aimed at personalization of management. World psychiatry : Official Journal of the World Psychiatric Association (WPA) 2021;20(1):4–33. doi: https://doi.org/10.1002/wps.20809

37. Martinez-Hernaez A, Pie-Balaguer A, Serrano-Miguel M, et al. The collaborative management of antipsychotic medication and its obstacles: A qualitative study. Social science & medicine (1982) 2020;247:112811. doi: https://doi.org/10.1016/j.socscimed.2020.112811

38. Yeisen RAH, Bjornestad J, Joa I, et al. Experiences of antipsychotic use in patients with early psychosis: a two-year follow-up study. BMC Psychiatry 2017;17(1):299. doi: https://doi.org/10.1186/s12888-017-1425-9

39. Younas M, Bradley E, Holmes N, et al. Mental health pharmacists views on shared decision-making for antipsychotics in serious mental illness. International Journal of Clinical Pharmacy 2016;38(5):1191–9. doi: https://doi.org/10.1007/s11096-016-0352-z

40. Geyt GL, Awenat Y, Tai S, et al. Personal Accounts of Discontinuing Neuroleptic Medication for Psychosis. Qualitative Health Research 2017;27(4):559–72. doi: https://doi.org/10.1177/1049732316634047

41. Grünwald LM, Thompson J. Re-starting the conversation: improving shared decision making in antipsychotic prescribing. Psychosis 2021;13(4):373–77. doi: https://doi.org/10.1080/17522439.2021.1903979

42. Grunwald LM, Duddy C, Byng R, et al. The role of trust and hope in antipsychotic medication reviews between GPs and service users a realist review. BMC Psychiatry 2021;21(1):390. doi: https://doi.org/10.1186/s12888-021-03355-3

43. Green CA, Polen MR, Janoff SL, et al. Understanding how clinician-patient relationships and relational continuity of care affect recovery from serious mental illness: STARS study results. Psychiatric Rehabilitation Journal 2008;32(1):9. doi: https://doi.org/10.2975/32.1.2008.9.22

44. Kaar SJ, Gobjila C, Butler E, et al. Making decisions about antipsychotics: a qualitative study of patient experience and the development of a decision aid. BMC Psychiatry 2019;19(1):309. doi: https://doi.org/10.1186/s12888-019-2304-3

45. Weiden PJ. Redefining Medication Adherence in the Treatment of Schizophrenia: How Current Approaches to Adherence Lead to Misinformation and Threaten Therapeutic Relationships. Psychiatric Clinics of North America 2016;39(2):199–216. doi: https://doi.org/10.1016/j.psc.2016.01.004

46. Aref-Adib G, O’Hanlon P, Fullarton K, et al. A qualitative study of online mental health information seeking behaviour by those with psychosis. BMC Psychiatry 2016;16:232. doi: https://doi.org/10.1186/s12888-016-0952-0

47. Crellin NE, Priebe S, Morant N, et al. An analysis of views about supported reduction or discontinuation of antipsychotic treatment among people with schizophrenia and other psychotic disorders. BMC Psychiatry 2022;22(1):1–11. doi: https://doi.org/10.1186/s12888-022-03822-5

48. Delman J, Clark JA, Eisen SV, et al. Facilitators and barriers to the active participation of clients with serious mental illnesses in medication decision making: the perceptions of young adult clients. The Journal of Behavioral Health Services & Research 2015;42(2):238–53. doi: https://doi.org/10.1007/s11414-014-9431-x

49. Kaminskiy E, Zisman-Ilani Y, Morant N, et al. Barriers and enablers to shared decision making in psychiatric medication management: A qualitative investigation of clinician and service users’ views. Frontiers in Psychiatry 2021;12:678005. doi: https://doi.org/10.3389/fpsyt.2021.678005

50. Katz S, Goldblatt H, Hasson-Ohayon I, et al. Retrospective accounts of the process of using and discontinuing psychiatric medication. Qualitative Health Research 2019;29(2):198–210. doi: https://doi.org/10.1177/1049732318793418

51. Keogh B, Murphy E, Doyle L, et al. Mental health service users experiences of medication discontinuation: a systematic review of qualitative studies. Journal of Mental Health 2022;31(2):227–38. doi: https://doi.org/10.1080/09638237.2021.1922644

52. King SR, Allan M, Lindsey L. “I found hundreds of other people… but I still wasn’t believed”–An exploratory study on lived experiences of antipsychotic withdrawal. Psychosis 2022:1–13. doi: https://doi.org/10.1080/17522439.2022.2141841

53. Opler LA, Ramirez PM, Dominguez LM, et al. Rethinking medication prescribing practices in an inner-city Hispanic mental health clinic. Journal of Psychiatric Practice 2004;10(2):134–40.

54. Pedley R, McWilliams C, Lovell K, et al. Qualitative systematic review of barriers and facilitators to patient-involved antipsychotic prescribing. BJPsych Open 2018;4(1):5–14. doi: https://doi.org/doi:10.1192/bjo.2017.5

55. Thompson J, Stansfeld JL, Cooper RE, et al. Experiences of taking neuroleptic medication and impacts on symptoms, sense of self and agency: a systematic review and thematic synthesis of qualitative data. Social Psychiatry and Psychiatric Epidemiology 2020;55(2):151–64. doi: https://doi.org/10.1007/s00127-019-01819-2

56. Larsen-Barr M, Seymour F, Read J, et al. Attempting to discontinue antipsychotic medication: Withdrawal methods, relapse and success. Psychiatry Research 2018;270:365–74. doi: https://doi.org/10.1016/j.psychres.2018.10.001

57. Bjornestad J, Lavik KO, Davidson L, et al. Antipsychotic treatment – a systematic literature review and meta-analysis of qualitative studies. Journal of Mental Health 2020;29(5):513–23. doi: https://doi.org/10.1080/09638237.2019.1581352

58. Clifford L, Crabb S, Turnbull D, et al. A qualitative study of medication adherence amongst people with schizophrenia. Archives of Psychiatric Nursing 2020;34(4):194–99. doi: https://doi.org/10.1016/j.apnu.2020.06.002

59. Haugom EW, Stensrud B, Beston G, et al. Mental health professionals’ experiences with shared decision-making for patients with psychotic disorders: a qualitative study. BMC Health Services Research 2020;20(1):1093. doi: https://doi.org/10.1186/s12913-020-05949-1

60. Jakovljevic M. Long-acting injectable (depot) antipsychotics and changing treatment philosophy: possible contribution to integrative care and personal recovery of schizophrenia. Psychiatria Danubina 2014;26(4):304–07.

61. Pietrini F, Albert U, Ballerini A, et al. The modern perspective for long-acting injectables antipsychotics in the patient-centered care of schizophrenia. Neuropsychiatric Disease and Treatment 2019;15:1045–60. doi: http://doi.org/10.2147/NDT.S199048

62. Salzmann-Erikson M, Sjodin M. A narrative meta-synthesis of how people with schizophrenia experience facilitators and barriers in using antipsychotic medication: Implications for healthcare professionals. International Journal of Nursing Studies 2018;85:7–18. doi: https://doi.org/10.1016/j.ijnurstu.2018.05.003

63. Velligan DI, Sajatovic M, Hatch A, et al. Why do psychiatric patients stop antipsychotic medication? A systematic review of reasons for nonadherence to medication in patients with serious mental illness. Patient Preference and Adherence 2017;11:449–68. doi: https://doi.org/10.2147/PPA.S124658

64. Watts M, Murphy E, Keogh B, et al. Deciding to discontinue prescribed psychotropic medication: A qualitative study of service users’ experiences. International Journal of Mental Health Nursing 2021;30:1395–406. doi: https://doi.org/10.1111/inm.12894

65. Fiore G, Bertani DE, Marchi M, et al. Patient subjective experience of treatment with long-acting injectable antipsychotics: a systematic review of qualitative studies. Jornal Brasileiro de Psiquiatria 2021;70:68–77. doi: https://doi.org/10.1590/0047-2085000000311

66. Haddad PM, Brain C, Scott J. Nonadherence with antipsychotic medication in schizophrenia: challenges and management strategies. Patient Related Outcome Measures 2014;5:43–62. doi: https://doi.org/10.2147/PROM.S42735

67. Matthias MS, Salyers MP, Rollins AL, et al. Decision making in recovery-oriented mental health care. Psychiatric rehabilitation journal 2012;35(4):305. doi: https://doi.org/10.2975/35.4.2012.305.314

68. Morant N, Azam K, Johnson S, et al. The least worst option: user experiences of antipsychotic medication and lack of involvement in medication decisions in a UK community sample. Journal of Mental Health 2018;27(4):322–28. doi: https://doi.org/10.1080/09638237.2017.1370637

69. Phan SV. Medication adherence in patients with schizophrenia. International Journal of Psychiatry in Medicine 2016;51(2):211–9. doi: https://doi.org/10.1177/0091217416636601

70. Rungruangsiripan M, Sitthimongkol Y, Maneesriwongul W, et al. Mediating role of illness representation among social support, therapeutic alliance, experience of medication side effects, and medication adherence in persons with schizophrenia. Archives of Psychiatric Nursing 2011;25(4):269–83. doi: https://doi.org/10.1016/j.apnu.2010.09.002

71. Seale C, Chaplin R, Lelliott P, et al. Sharing decisions in consultations involving anti-psychotic medication: a qualitative study of psychiatrists’ experiences. Social Science & Medicine 2006;62(11):2861–73. doi: https://doi.org/10.1016/j.socscimed.2005.11.002

72. Mahone IH, Maphis CF, Snow DE. Effective Strategies for Nurses Empowering Clients With Schizophrenia: Medication Use as a Tool in Recovery. Issues in Mental Health Nursing 2016;37(5):372–9. doi: https://doi.org/10.3109/01612840.2016.1157228

73. Alguera-Lara V, Dowsey MM, Ride J, et al. Shared decision making in mental health: the importance for current clinical practice. Australasian psychiatry : Bulletin of Royal Australian and New Zealand College of Psychiatrists 2017;25(6):578–82. doi: https://doi.org/10.1177/1039856217734711

74. Fiorillo A, Barlati S, Bellomo A, et al. The role of shared decision-making in improving adherence to pharmacological treatments in patients with schizophrenia: a clinical review. Annals of general psychiatry 2020;19:43. doi: https://doi.org/10.1186/s12991-020-00293-4

75. Hansen H, Stige SH, Davidson L, et al. How do people experience early intervention services for psychosis? A meta-synthesis. Qualitative Health Research 2018;28(2):259–72. doi: https://doi.org/10.1177/1049732317735080

76. Jawad I, Watson S, Haddad PM, et al. Medication nonadherence in bipolar disorder: a narrative review. Therapeutic Advances in Psychopharmacology 2018;8(12):349–63. doi: https://doi.org/10.1177/2045125318804364

77. Leclerc E, Noto C, Bressan RA, et al. Determinants of adherence to treatment in first-episode psychosis: a comprehensive review. Revista Brasileira de Psiquiatria 2015;37(2):168–76. doi: https://doi.org/10.1590/1516-4446-2014-1539

78. Lim M, Li Z, Xie H, et al. The Effect of Therapeutic Alliance on Attitudes Toward Psychiatric Medications in Schizophrenia. Journal of Clinical Psychopharmacology 2021;41(5):551–60. doi: https://doi.org/10.1097/JCP.0000000000001449

79. Pinfold V, Dare C, Hamilton S, et al. Anti-psychotic medication decision making during pregnancy: A co-produced research study. Mental Health Review Journal 2019;24(2):69–84. doi: https://doi.org/10.1108/MHRJ-04-2017-0018

80. Quirk A, Chaplin R, Hamilton S, et al. Communication about adherence to long-term antipsychotic prescribing: an observational study of psychiatric practice. Social Psychiatry and Psychiatric Epidemiology 2013;48:639–47. doi: https://doi.org/10.1007/s00127-012-0581-y

81. Sowerby C, Taylor D. Cross-sector user and provider perceptions on experiences of shared-care clozapine: a qualitative study. BMJ Open 2017;7(9):e017183. doi: (https://doi.org/10.1136/bmjopen-2017-017183

82. Garcia S, Martinez-Cengotitabengoa M, Lopez-Zurbano S, et al. Adherence to Antipsychotic Medication in Bipolar Disorder and Schizophrenic Patients: A Systematic Review. Journal of Clinical Psychopharmacology 2016;36(4):355–71. doi: https://doi.org/10.1097/JCP.0000000000000523

83. Steingard S. Five year outcomes of tapering antipsychotic drug doses in a community mental health center. Community Mental Health Journal 2018;54(8):1097–100. doi: https://doi.org/10.1007/s10597-018-0313-1

84. Roberts R, Neasham A, Lambrinudi C, et al. A thematic analysis assessing clinical decision-making in antipsychotic prescribing for schizophrenia. BMC Psychiatry 2018;18(1):290. doi: https://doi.org/10.1186/s12888-018-1872-y

85. Zisman-Ilani Y, Lysaker PH, Hasson-Ohayon I. Shared Risk Taking: Shared Decision Making in Serious Mental Illness. Psychiatric Services 2021;72(4):461–63. doi: https://doi.org/10.1176/appi.ps.202000156

86. Larsen-Barr M, Seymour F. Service-user efforts to maintain their wellbeing during and after successful withdrawal from antipsychotic medication. Therapeutic Advances in Psychopharmacology 2021;11:2045125321989133. doi: https://doi.org/10.1177/2045125321989133

87. Lewins A, Morant N, Akther-Robertson J, et al. A qualitative exploration of family members’ perspectives on reducing and discontinuing antipsychotic medication. Journal of Mental Health 2022:1–8. doi: https://doi.org/10.1080/09638237.2022.2069710

88. Zipursky RB, Odejayi G, Agid O, et al. You say “schizophrenia” and I say “psychosis”: just tell me when I can come off this medication. Schizophrenia Research 2020;225:39–46. doi: https://doi.org/10.1016/j.schres.2020.02.009

89. Ostrow L, Croft B, Weaver A, et al. An exploratory analysis of the role of social supports in psychiatric medication discontinuation: results related to family involvement. Psychosis 2019;11(3):212–22. doi: https://doi.org/10.1080/17522439.2019.1615110

90. Zhou Y, Rosenheck R, Mohamed S, et al. Factors associated with complete discontinuation of medication among patients with schizophrenia in the year after hospital discharge. Psychiatry Research 2017;250:129–35. doi: https://doi.org/10.1016/j.psychres.2017.01.036

91. Coulthard K, Patel D, Brizzolara C, et al. A feasibility study of expert patient and community mental health team led bipolar psychoeducation groups: implementing an evidence based practice. BMC psychiatry 2013;13(1):1–12. doi: https://doi.org/10.1186/1471-244X-13-301

92. Evans M, Barker H, Peddireddy S, et al. Peer-delivered services and peer support reaching people with schizophrenia: A scoping review. International Journal of Mental Health 2021:1–23. doi: https://doi.org/10.1080/00207411.2021.1975441

93. Rossiter C, Levett-Jones T, Pich J. The impact of person-centred care on patient safety: An umbrella review of systematic reviews. International Journal of Nursing Studies 2020;109:103658. doi: https://doi.org/10.1016/j.ijnurstu.2020.103658

94. Davidson G, Kelly B, Macdonald G, et al. Supported decision making: A review of the international literature. International Journal of Law and Psychiatry 2015;38:61–67. doi: https://doi.org/10.1016/j.ijlp.2015.01.008

95. Knight F, Kokanović R, Ridge D, et al. Supported Decision-Making: The Expectations Held by People With Experience of Mental Illness. Qualitative Health Research 2018;28(6):1002–15. doi: https://doi.org/10.1177/1049732318762371

96. Kokanović R, Brophy L, McSherry B, et al. Supported decision-making from the perspectives of mental health service users, family members supporting them and mental health practitioners. Australian & New Zealand Journal of Psychiatry 2018;52(9):826–33. doi: https://doi.org/10.1177/0004867418784177

97. Penzenstadler L, Molodynski A, Khazaal Y. Supported decision making for people with mental health disorders in clinical practice: a systematic review. International Journal of Psychiatry in Clinical Practice 2020;24(1):3–9. doi: https://doi.org/10.1080/13651501.2019.1676452

98. Lewicki RJ, Tomlinson EC, Gillespie N. Models of interpersonal trust development: Theoretical approaches, empirical evidence, and future directions. Journal of Management 2006;32(6):991–1022. doi: https://doi.org/10.1177/0149206306294405

99. Maidment ID, Brown P, Calnan M. An exploratory study of the role of trust in medication management within mental health services. International Journal of Clinical Pharmacy 2011;33(4):614–20. doi: https://doi.org/10.1007/s11096-011-9510-5

100. Brown P, Calnan M. Trust as a Means of Bridging the Management of Risk and the Meeting of Need: A Case Study in Mental Health Service Provision. Social Policy & Administration 2013;47(3):242–61. doi: https://doi.org/10.1111/j.1467-9515.2012.00865.x

101. Brown P, Calnan M, Scrivener A, et al. Trust in Mental Health Services: A neglected concept. Journal of Mental Health 2009;18(5):449–58. doi: https://doi.org/10.3109/09638230903111122

102. Trusty WT, Penix EA, Dimmick AA, et al. Shared decision-making in mental and behavioural health interventions. Journal of Evaluation in Clinical Practice 2019;25(6):1210–16. doi: https://doi.org/10.1111/jep.13255

103. Ahmed N, Barlow S, Reynolds L, et al. Mental health professionals’ perceived barriers and enablers to shared decision-making in risk assessment and risk management: a qualitative systematic review. BMC Psychiatry 2021;21(1):594. doi: https://doi.org/10.1186/s12888-021-03304-0

104. Markham S. Collaborative risk assessment in secure and forensic mental health settings in the UK. General Psychiatry 2020;33(5):e100291. doi: https://doi.org/10.1136/gpsych-2020-100291

105. Department of Health. Best practice in managing risk: principles and evidence for practice in the assessment and management of risk to self and others in mental health services. London, UK, 2009.

106. Shalaby RAH, Agyapong VIO. Peer Support in Mental Health: Literature Review. JMIR Ment Health 2020;7(6):e15572. doi: https://doi.org/10.2196/15572

107. Thomas EC, Ben-David S, Treichler E, et al. A Systematic Review of Shared Decision–Making Interventions for Service Users With Serious Mental Illnesses: State of the Science and Future Directions. Psychiatric Services 2021;72(11):1288–300. doi: https://doi.org/10.1176/appi.ps.202000429

108. Kapadia D, Zhang J, Salway S, et al. Ethnic Inequalities in Healthcare: A Rapid Evidence Review: NHS Race & Health Observatory 2022.

109. Bahorik AL, Satre DD, Kline-Simon AH, et al. Serious mental illness and medical comorbidities: Findings from an integrated health care system. Journal of Psychosomatic Research 2017;100:35–45. doi: https://doi.org/10.1016/j.jpsychores.2017.07.004

110. Rodgers M, Dalton J, Harden M, et al. Integrated Care to Address the Physical Health Needs of People with Severe Mental Illness: A Mapping Review of the Recent Evidence on Barriers, Facilitators and Evaluations. Int J Integr Care 2018;18(1):9. doi: https://doi.org/10.5334/ijic.2605

