## Supplemental File 1, and will be used for the link to the file on the preprint slide for "A realist review of medication optimisation of community dwelling service users with serious mental illness"

### Supplementary File 1: MEDIATE search strategies

#### Main database search

##### MEDLINE

Host: Ovid

Data parameters: Ovid MEDLINE® ALL

Date range searched: 1946 to present (Daily update)

Date searched: 12/01/2022

Searcher: CD

Hits: n=434

| 1 | (serious mental illness* or serious mental disorder*).ti,ab,kw. | 4786 |
| --- | --- | --- |
| 2 | (severe mental illness* or severe mental disorder*).ti,ab,kw. | 6762 |
| 3 | SMI.ti,ab,kw. | 5722 |
| 4 | (schizophren* or schizoaffective or psychosis or psychotic or bipolar disorder* or personality disorder*).ti,ab,kw. | 214792 |
| 5 | exp *"bipolar and related disorders"/ or exp *"schizophrenia spectrum and other psychotic disorders"/ or exp *"personality disorders"/ | 183862 |
| 6 | or/1-5 | 274623 |
| 7 | (antipsychotic* or anti-psychotic* or psychotropic* or neuroleptic*).ti,ab,kw. | 76358 |
| 8 | (clozapine or olanzapine or quetiapine or risperidone or aripiprazole or haloperidol).ti,ab,kw. | 43567 |
| 9 | exp *Antipsychotic Agents/ | 79470 |
| 10 | ((medication or medicine*) adj1 (optimi* or manag*)).ti,ab,kw. | 5678 |
| 11 | ((appropriate* or inappropriate*) adj2 prescri*).ti,ab,kw. | 5669 |
| 12 | (deprescri* OR de-prescri*).ti,ab,kw. | 351 |
| 13 | (polypharmacy or poly-pharmacy).ti,ab,kw. | 9476 |
| 14 | *Medication Therapy Management/ | 1596 |
| 15 | *deprescriptions/ | 571 |
| 16 | Inappropriate Prescribing/ | 4146 |
| 17 | or/7-16 | 158205 |
| 18 | ((shared or joint or collab*) adj1 decision*).ti,ab,kw. | 11704 |
| 19 | *Decision Making, Shared/ | 764 |
| 20 | ((decision or communication) adj1 (aid* or support* or tool*)).ti,ab,kw. | 27810 |
| 21 | *decision support techniques/ | 12771 |
| 22 | or/18-21 | 47905 |
| 23 | ((patient* or client* or service user* or person or people) adj1 (centred or centered or participation or involvement or engagement or preferen* or empower* or self-determin*)).ti,ab,kw. | 60459 |
| 24 | *Patient Participation/ | 16117 |
| 25 | exp *Patient-Centered Care/ | 14278 |
| 26 | or/23-25 | 79410 |
| 27 | ((clinician* or doctor* or physician* or psychiatrist* or psychologist or nurse* or pharmacist* or health care professional* or healthcare professional* or social care professional* or prescriber*) adj1 (patient* or client* or service user* or family or carer*) adj1 (relations* or communic* or cooperat* or partnership* or collab*)).ti,ab,kw. | 14629 |
| 28 | therapeutic alliance.ti,ab,kw. | 2919 |
| 29 | *professional-family relations/ or *professional-patient relations/ or *nurse-patient relations/ or *physician-patient relations/ or *therapeutic alliance/ | 73250 |
| 30 | or/27-29 | 83679 |
| 31 | 6 and 17 and (22 or 26 or 30) | 475 |
| 32 | limit 31 to english language | 434 |

##### Embase

Host: Ovid

Data parameters: Embase 1974 to present

Date range searched: 1974 to present (Daily update)

Date searched: 12/01/2022

Searcher: CD

Hits: n=615

| 1 | (serious mental illness* or serious mental disorder*).ti,ab,kw. | 5844 |
| --- | --- | --- |
| 2 | (severe mental illness* or severe mental disorder*).ti,ab,kw. | 9002 |
| 3 | SMI.ti,ab,kw. | 8497 |
| 4 | (schizophren* or schizoaffective or psychosis or psychotic or bipolar disorder* or personality disorder*).ti,ab,kw. | 288867 |
| 5 | exp *bipolar disorder/ or exp *schizophrenia/ or exp *psychosis/ or exp *personality disorder/ | 232560 |
| 6 | or/1-5 | 347444 |
| 7 | (antipsychotic* or anti-psychotic* or psychotropic* or neuroleptic*).ti,ab,kw. | 111542 |
| 8 | (clozapine or olanzapine or quetiapine or risperidone or aripiprazole or haloperidol).ti,ab,kw. | 61242 |
| 9 | exp *neuroleptic agent/ | 129929 |
| 10 | ((medication or medicine*) adj1 (optimi* or manag*)).ti,ab,kw. | 9631 |
| 11 | ((appropriate* or inappropriate*) adj2 prescri*).ti,ab,kw. | 9644 |
| 12 | (deprescri* OR de-prescri*).ti,ab,kw. | 2065 |
| 13 | (polypharmacy or poly-pharmacy).ti,ab,kw. | 16197 |
| 14 | *Medication Therapy Management/ | 4868 |
| 15 | *deprescription/ or *polypharmacy/ | 5340 |
| 16 | exp *inappropriate prescribing/ | 2850 |
| 17 | or/7-16 | 238346 |
| 18 | ((shared or joint or collab*) adj1 decision*).ti,ab,kw. | 16561 |
| 19 | *shared decision making/ | 2939 |
| 20 | ((decision or communication) adj1 (aid* or support* or tool*)).ti,ab,kw. | 37538 |
| 21 | *decision support system/ | 11336 |
| 22 | or/18-21 | 57329 |
| 23 | ((patient* or client* or service user* or person or people) adj1 (centred or centered or participation or involvement or engagement or preferen* or empower* or self-determin*)).ti,ab,kw. | 86326 |
| 24 | *Patient Participation/ | 10500 |
| 25 | or/23-24 | 94070 |
| 26 | ((clinician* or doctor* or physician* or psychiatrist* or psychologist or nurse* or pharmacist* or health care professional* or healthcare professional* or social care professional* or prescriber*) adj1 (patient* or client* or service user* or family or carer*) adj1 (relations* or communic* or cooperat* or partnership* or collab*)).ti,ab,kw. | 18113 |
| 27 | therapeutic alliance.ti,ab,kw. | 4206 |
| 28 | exp *professional-patient relationship/ | 20671 |
| 29 | or/26-28 | 41129 |
| 30 | 6 and 17 and (22 or 25 or 29) | 668 |
| 31 | limit 30 to english language | 615 |

##### PsycINFO

Host: Ovid

Data parameters: PsycINFO 1806 to present

Date range searched: 1806 to present (Weekly update)

Date searched: 12/01/2022

Searcher: CD

Hits: n=286

| 1 | (serious mental illness* or serious mental disorder*).ti,ab. | 5512 |
| --- | --- | --- |
| 2 | (severe mental illness* or severe mental disorder*).ti,ab. | 6876 |
| 3 | SMI.ti,ab. | 2493 |
| 4 | (schizophren* or schizoaffective or psychosis or psychotic or bipolar disorder* or personality disorder*).ti,ab. | 215283 |
| 5 | exp *psychosis/ or exp *acute psychosis/ or exp *chronic psychosis/ or exp *"paranoia (psychosis)"/ or exp *schizophrenia/ or exp *personality disorders/ or exp *bipolar disorder/ | 159102 |
| 6 | or/1-5 | 241238 |
| 7 | (antipsychotic* or anti-psychotic* or psychotropic* or neuroleptic*).ti,ab. | 51054 |
| 8 | (clozapine or olanzapine or quetiapine or risperidone or aripiprazole or haloperidol).ti,ab. | 25798 |
| 9 | exp *neuroleptic drugs/ | 28609 |
| 10 | ((medication or medicine*) adj1 (optimi* or manag*)).ti,ab. | 1893 |
| 11 | ((appropriate* or inappropriate*) adj2 prescri*).ti,ab. | 735 |
| 12 | (deprescrib* or de-prescrib*).ti,ab. | 126 |
| 13 | (polypharmacy or poly-pharmacy).ti,ab. | 2180 |
| 14 | *polypharmacy/ | 1005 |
| 15 | or/7-14 | 67584 |
| 16 | ((shared or joint or collab*) adj1 decision*).ti,ab. | 3880 |
| 17 | ((decision or communication) adj1 (aid* or support* or tool*)).ti,ab. | 8348 |
| 18 | *decision support systems/ | 3185 |
| 19 | or/16-18 | 13413 |
| 20 | ((patient* or client* or service user* or person or people) adj1 (centred or centered or participation or involvement or engagement or preferen* or empower* or self-determin*)).ti,ab. | 24559 |
| 21 | *client participation/ | 2033 |
| 22 | *patient centered care/ | 242 |
| 23 | or/20-22 | 25793 |
| 24 | ((clinician* or doctor* or physician* or psychiatrist* or psychologist or nurse* or pharmacist* or health care professional* or healthcare professional* or social care professional* or prescriber*) adj1 (patient* or client* or service user* or family or carer*) adj1 (relations* or communic* or cooperat* or partnership* or collab*)).ti,ab. | 6086 |
| 25 | therapeutic alliance.ti,ab. | 5475 |
| 26 | *therapeutic alliance/ | 4507 |
| 27 | or/24-26 | 13757 |
| 28 | 6 and 15 and (19 or 23 or 27) | 325 |
| 29 | limit 28 to english language | 286 |

##### CINAHL (Cumulative Index to Nursing and Allied Health Literature)

Host: EbscoHOST

Data parameters: CINAHL 1981 onwards

Date range searched: 1981 to present (Update unknown)

Date searched: 12/01/2022

Searcher: CD

Hits: n=279

| S32 | S6 AND S17 AND S30 | 279 |
| --- | --- | --- |
| S31 | S6 AND S17 AND S30 | 282 |
| S30 | S21 OR S25 OR S29 | 147,278 |
| S29 | S26 OR S27 OR S28 | 61,351 |
| S28 | (MM "Professional-Client Relations+") OR (MM "Professional-Patient Relations+") OR (MM "Professional-Family Relations") | 54,797 |
| S27 | TI "therapeutic alliance" OR AB "therapeutic alliance" | 1,531 |
| S26 | TI ( (clinician* or doctor* or physician* or psychiatrist* or psychologist or nurse* or pharmacist* or "health care professional*" or "healthcare professional*" or "social care professional*" or prescriber*) N1 (patient* or client* or "service user*" or family or carer*) N1 (relations* or communic* or cooperat* or partnership* or collab*) ) OR AB ( (clinician* or doctor* or physician* or psychiatrist* or psychologist or nurse* or pharmacist* or "health care professional*" or "healthcare professional*" or "social care professional*" or prescriber*) N1 (patient* or client* or "service user*" or family or carer*) N1 (relations* or communic* or cooperat* or partnership* or collab*) ) | 9,412 |
| S25 | S22 OR S23 OR S24 | 67,636 |
| S24 | (MM "Patient Centered Care") | 15,994 |
| S23 | (MM "Consumer Participation") | 12,845 |
| S22 | TI ( (patient* or client* or "service user*" or person or people) N1 (centred or centered or participation or involvement or engagement or preferen* or empower* or self-determin*) ) OR AB ( (patient* or client* or "service user*" or person or people) N1 (centred or centered or participation or involvement or engagement or preferen* or empower* or self-determin*) ) | 49,364 |
| S21 | S18 OR S19 OR S20 | 28,396 |
| S20 | TI ( (decision or communication) N1 (aid* or support* or tool*) ) OR AB ( (decision or communication) N1 (aid* or support* or tool*) ) | 18,961 |
| S19 | (MM "Decision Making, Shared") OR (MM "Decision Support Techniques") | 5,267 |
| S18 | TI ( (shared or joint or collab*) N1 decision* ) OR AB ( (shared or joint or collab*) N1 decision* ) | 7,568 |
| S17 | S7 OR S8 OR S9 OR S10 OR S11 OR S12 OR S13 OR S14 OR S15 OR S16 | 42,399 |
| S16 | (MM "Inappropriate Prescribing") | 2,032 |
| S15 | (MM "Polypharmacy+") | 2,311 |
| S14 | (MM "Medication Management") | 642 |
| S13 | TI ( polypharmacy or poly-pharmacy ) OR AB ( polypharmacy or poly-pharmacy ) | 4,234 |
| S12 | TI ( deprescri* OR de-prescri* ) OR AB ( deprescri* OR de-prescri* ) | 1,054 |
| S11 | TI ( (appropriate* or inappropriate*) N2 prescri* ) OR AB ( (appropriate* or inappropriate*) N2 prescri* ) | 3,228 |
| S10 | TI ( (medication or medicine*) N1 (optimi* or manag*) ) OR AB ( (medication or medicine*) N1 (optimi* or manag*) ) | 6,678 |
| S9 | (MM "Antipsychotic Agents+") | 14,028 |
| S8 | TI ( clozapine or olanzapine or quetiapine or risperidone or aripiprazole or haloperidol ) OR AB ( clozapine or olanzapine or quetiapine or risperidone or aripiprazole or haloperidol ) | 8,735 |
| S7 | TI ( antipsychotic* OR anti-psychotic* OR psychotropic* OR neuroleptic* ) OR AB ( antipsychotic* OR anti-psychotic* OR psychotropic* OR neuroleptic* ) | 18,990 |
| S6 | S1 OR S2 OR S3 OR S4 OR S5 | 141,494 |
| S5 | (MM "Psychotic Disorders+") OR (MM "Personality Disorders+") | 117,057 |
| S4 | TI ( schizophren* OR schizoaffective OR psychosis OR psychotic OR "bipolar disorder*" OR "personality disorder*" ) OR AB ( schizophren* OR schizoaffective OR psychosis OR psychotic OR "bipolar disorder*" OR "personality disorder*" ) | 57,164 |
| S3 | TI SMI OR AB SMI | 2,222 |
| S2 | TI ( "severe mental illness*" OR "severe mental disorder*" ) OR AB ( "severe mental illness*" OR "severe mental disorder*" ) | 3,684 |
| S1 | TI ( "serious mental illness*" OR "serious mental disorder*" ) OR AB ( "serious mental illness*" OR "serious mental disorder*" ) | 3,376 |

##### Cochrane Library

Host: Cochrane Library

Data parameters: CDSR, Protocols, CENTRAL (trials), Editorials, Special Collections, Clinical Answers

Date range searched: No limit

Date searched: 12/01/2022

Searcher: CD

Hits: n=164

| #1 | ("serious mental illness*" or "serious mental disorder*"):ti,ab,kw | 739 |
| --- | --- | --- |
| #2 | ("severe mental illness*" or "severe mental disorder*"):ti,ab,kw | 1005 |
| #3 | (SMI):ti,ab,kw | 839 |
| #4 | (schizophren* or schizoaffective or psychosis or psychotic or "bipolar disorder*" or "personality disorder*"):ti,ab,kw | 30237 |
| #5 | MeSH descriptor: [Bipolar and Related Disorders] explode all trees | 2813 |
| #6 | MeSH descriptor: [Schizophrenia Spectrum and Other Psychotic Disorders] explode all trees | 9647 |
| #7 | MeSH descriptor: [Personality Disorders] explode all trees | 1458 |
| #8 | #1 OR #2 OR #3 OR #4 OR #5 OR #6 OR #7 | 31816 |
| #9 | *(antipsychotic* or anti-psychotic* or psychotropic* or neuroleptic*):ti,ab,kw* | 14078 |
| #10 | (clozapine or olanzapine or quetiapine or risperidone or aripiprazole or haloperidol):ti,ab,kw | 10945 |
| #11 | MeSH descriptor: [Antipsychotic Agents] explode all trees | 4774 |
| #12 | (((medication or medicine*) near/1 (optimi* or manag*))):ti,ab,kw | 1166 |
| #13 | (((appropriate* or inappropriate*) near/2 prescri*)):ti,ab,kw | 842 |
| #14 | ((deprescri* OR de-prescri*)):ti,ab,kw | 301 |
| #15 | ((polypharmacy or poly-pharmacy)):ti,ab,kw | 1151 |
| #16 | MeSH descriptor: [undefined] explode all trees | 0 |
| #17 | MeSH descriptor: [Deprescriptions] this term only | 43 |
| #18 | MeSH descriptor: [Inappropriate Prescribing] this term only | 169 |
| #19 | #9 OR #10 OR #11 OR #12 OR #13 OR #14 OR #15 OR #16 OR #17 OR #18 | 22129 |
| #20 | (((shared or joint or collab*) near/1 decision*)):ti,ab,kw | 1730 |
| #21 | MeSH descriptor: [Decision Making, Shared] this term only | 65 |
| #22 | (((decision or communication) near/1 (aid* or support* or tool*))):ti,ab,kw | 5107 |
| #23 | MeSH descriptor: [Decision Support Techniques] this term only | 881 |
| #24 | #20 OR #21 OR #22 OR #23 | 6275 |
| #25 | (((patient* or client* or "service user*" or person or people) near/1 (centred or centered or participation or involvement or engagement or preferen* or empower* or self-determin*))):ti,ab,kw | 14309 |
| #26 | MeSH descriptor: [Patient Participation] this term only | 1489 |
| #27 | MeSH descriptor: [Patient-Centered Care] explode all trees | 813 |
| #28 | #25 OR #26 OR #27 | 14429 |
| #29 | (((clinician* or doctor* or physician* or psychiatrist* or psychologist or nurse* or pharmacist* or "health care professional*" or "healthcare professional*" or "social care professional*" or prescriber*) near/1 (patient* or client* or "service user*" or family or carer*) near/1 (relations* or communic* or cooperat* or partnership* or collab*))):ti,ab,kw | 2791 |
| #30 | ("therapeutic alliance"):ti,ab,kw | 787 |
| #31 | MeSH descriptor: [undefined] explode all trees | 0 |
| #32 | MeSH descriptor: [Professional-Patient Relations] this term only | 792 |
| #33 | MeSH descriptor: [Nurse-Patient Relations] this term only | 397 |
| #34 | MeSH descriptor: [Physician-Patient Relations] this term only | 1456 |
| #35 | MeSH descriptor: [Therapeutic Alliance] this term only | 53 |
| #36 | #29 OR #30 OR #31 OR #32 OR #33 OR #34 OR #35 | 4205 |
| #37 | #24 OR #28 OR #36 | 22884 |
| #38 | #8 AND #19 AND #37 | 164 |

##### Scopus

Host: Scopus.com

Data parameters: n/a

Date range searched: No limit

Date searched: 13/01/2022

Searcher: CD

Hits: n=368

| Full string | ( TITLE-ABS ( "serious mental illness*" OR "serious mental disorder*" OR "severe mental illness*" OR "severe mental disorder*" OR smi OR schizophren* OR schizoaffective OR psychosis OR psychotic OR "bipolar disorder*" OR "personality disorder*" ) ) AND ( ( TITLE-ABS ( antipsychotic OR anti-psychotic* OR psychotropic* OR neuroleptic* OR clozapine OR olanzapine OR quetiapine OR risperidone OR aripiprazole OR haloperidol ) ) OR ( TITLE-ABS ( ( medication OR medicine ) W/1 ( optimi* OR manag* ) ) ) OR ( TITLE-ABS ( ( appropriate* OR inappropriate* ) W/2 ( prescri* ) ) ) OR ( TITLE-ABS ( deprescri* OR de-prescri* ) ) OR ( TITLE-ABS ( polypharmacy OR poly-pharmacy ) ) ) AND ( ( TITLE-ABS ( ( shared OR joint OR collab* ) W/1 decision* ) ) OR ( TITLE-ABS ( ( decision OR communication ) W/1 ( aid* OR support* OR tool* ) ) ) OR ( TITLE-ABS ( ( patient* OR client* OR "service user*" OR person OR people ) W/1 ( centred OR centered OR participation OR involvement OR engagement OR preferen* OR empower* OR self-determin* ) ) ) OR ( TITLE-ABS ( ( clinician* OR doctor* OR physician* OR psychiatrist* OR psychologist OR nurse* OR pharmacist* OR "health care professional*" OR "healthcare professional*" OR "social care professional*" OR prescriber* ) W/1 ( patient* OR client* OR "service user*" OR family OR carer* ) W/1 ( relations* OR communic* OR cooperat* OR partnership* OR collab* ) ) ) OR ( TITLE-ABS ( "therapeutic alliance" ) ) ) AND ( LIMIT-TO ( LANGUAGE , "English" ) ) | 368 |
| --- | --- | --- |

##### Web of Science (Core)

Host: Web of Science (Clarivate Analytics)

Data parameters: SCIE, SSCI, SHCI, ESCI, CPCI, BKCI indexes

Date range searched: Unknown

Date searched: 13/01/2022

Searcher: CD

Hits: n=368

| 1 | TI=(("Serious mental illness*" OR "serious mental disorder*" OR "severe mental illness*" OR "severe mental disorder*" OR SMI OR schizophren* OR schizoaffective OR psychosis OR psychotic OR "bipolar disorder*" OR "personality disorder*")) OR AB=(("Serious mental illness*" OR "serious mental disorder*" OR "severe mental illness*" OR "severe mental disorder*" OR SMI OR schizophren* OR schizoaffective OR psychosis OR psychotic OR "bipolar disorder*" OR "personality disorder*")) | 388022 |
| --- | --- | --- |
| 2 | TI=(antipsychotic* OR anti-psychotic* OR psychotropic* OR neuroleptic* OR clozapine OR olanzapine OR quetiapine OR risperidone OR aripiprazole OR haloperidol) OR AB=(antipsychotic* OR anti-psychotic* OR psychotropic* OR neuroleptic* OR clozapine OR olanzapine OR quetiapine OR risperidone OR aripiprazole OR haloperidol) | 149751 |
| 3 | TI=(((medication OR Medicine*) Near/1 (optimi* OR manag*))) OR AB=(((medication OR Medicine*) Near/1 (optimi* OR manag*))) | 14929 |
| 4 | TI=(((appropriate* OR inappropriate*) near/2 prescri*)) OR AB=(((appropriate* OR inappropriate*) near/2 prescri*)) | 8944 |
| 5 | TI=((deprescri* OR de-prescri*)) OR AB=((deprescri* OR de-prescri*)) | 2905 |
| 6 | TI=((polypharmacy OR poly-pharmacy)) OR AB=((polypharmacy OR poly-pharmacy)) | 11643 |
| 7 | #2 OR #3 OR #4 OR #5 OR #6 | 183623 |
| 8 | TI=(((shared OR joint OR collab*) near/1 decision*)) OR AB=(((shared OR joint OR collab*) near/1 decision*)) | 19711 |
| 9 | TI=((decision OR communication) near/1 (aid* or support* or tool*)) OR AB=((decision OR communication) near/1 (aid* or support* or tool*)) | 154797 |
| 10 | TI=((patient* or client* or "service user*" or person or people) near/1 (centred or centered or participation or involvement or engagement or preferen* or empower* or self-determin*)) OR AB=((patient* or client* or "service user*" or person or people) near/1 (centred or centered or participation or involvement or engagement or preferen* or empower* or self-determin*)) | 138302 |
| 11 | TI=(((clinician* or doctor* or physician* or psychiatrist* or psychologist or nurse* or pharmacist* or "health care professional*" or "healthcare professional*" or "social care professional*" or prescriber*) near/1 (patient* or client* or "service user*" or family or carer*)) near/1 (relations* or communic* or cooperat* or partnership* or collab*)) OR AB=(((clinician* or doctor* or physician* or psychiatrist* or psychologist or nurse* or pharmacist* or "health care professional*" or "healthcare professional*" or "social care professional*" or prescriber*) near/1 (patient* or client* or "service user*" or family or carer*)) near/1 (relations* or communic* or cooperat* or partnership* or collab*)) | 21189 |
| 12 | TI=("therapeutic alliance") OR AB=("therapeutic alliance") | 4149 |
| 13 | #12 OR #11 OR #10 OR #9 OR #8 | 328533 |
| 14 | #13 AND #7 AND #1 | 483 |
| 15 | #13 AND #7 AND #1 and Web of Science Core Collection (Database) and English (Languages) | 368 |

##### Sociological Abstracts

Host: Proquest

Data parameters: 1952 to present (update unknown)

Date range searched: 1952 to present

Date searched: 13/01/2022

Searcher: CD

Hits: n=8

| Full string | (noft("serious mental illness*" OR "serious mental disorder*" OR "severe mental illness*" OR "severe mental disorder*" OR SMI) OR noft(schizophren* OR schizoaffective OR psychosis OR psychotic OR "bipolar disorder*" OR "personality disorder*")) AND (noft(antipsychotic* OR anti-psychotic* OR psychotropic* OR neuroleptic* OR clozapine OR olanzapine OR quetiapine or risperidone or aripiprazole or haloperidol) OR noft((medication or medicine*) N1 (optimi* or manag*)) OR noft((medication OR medicine*) NEAR/1 (optimi* OR manag*)) OR noft((appropriate* OR inappropriate*) N/2 (prescri*)) OR noft(deprescri* OR de-prescri*) OR noft(polypharmacy OR poly-pharmacy)) AND (noft((shared OR joint OR collab*) N/1 decision*) OR noft((decision OR communication) N/1 (aid* OR support* OR tool*)) OR noft((patient* or client* or ("service user" OR "service users") or person or people) N/1 (centred or centered or participation or involvement or engagement or preferen* or empower* or self-determin*)) OR noft((clinician* or doctor* or physician* or psychiatrist* or psychologist or nurse* or pharmacist* or "health care professional*" or ("healthcare professionals") or "social care professional*" or prescriber*) N/1 (patient* or client* or ("service user" OR "service users") or family or carer*) N/1 (relations* or communic* or cooperat* or partnership* or collab*)) OR noft("therapeutic alliance"))Limits applied | 8 |
| --- | --- | --- |

#### Additional search 1: Internet use for health information

##### MEDLINE

Host: Ovid

Data parameters: Ovid MEDLINE® ALL

Date range searched: 1946 to present (Daily update)

Date searched: 16/06/2022

Searcher: CD

Hits: n=51

| 1 | (serious mental illness* or serious mental disorder*).ti. | 1985 |
| --- | --- | --- |
| 2 | (severe mental illness* or severe mental disorder*).ti. | 2861 |
| 3 | SMI.ti. | 274 |
| 4 | (schizophren* or schizoaffective or psychosis or psychotic or bipolar disorder* or personality disorder*).ti. | 135488 |
| 5 | exp *"bipolar and related disorders"/ or exp *"schizophrenia spectrum and other psychotic disorders"/ or exp *"personality disorders"/ | 187573 |
| 6 | or/1-5 | 213216 |
| 7 | (internet or online or digital or web* or social media).ti. | 131598 |
| 8 | exp *Internet/ | 51669 |
| 9 | or/7-8 | 150150 |
| 10 | *Access to Information/ | 3801 |
| 11 | *Information Seeking Behavior/ | 1981 |
| 12 | *Patient Education as Topic/ | 40821 |
| 13 | exp *Health Education/ | 152838 |
| 14 | or/10-13 | 157860 |
| 15 | 6 and 9 and 14 | 51 |

##### PsycINFO

Host: Ovid

Data parameters: PsycINFO 1806 to present

Date range searched: 1806 to present (Weekly update)

Date searched: 16/06/2022

Searcher: CD

Hits: n=8

| 1 | (serious mental illness* or serious mental disorder*).ti. | 2052 |
| --- | --- | --- |
| 2 | (severe mental illness* or severe mental disorder*).ti. | 2622 |
| 3 | SMI.ti. | 66 |
| 4 | (schizophren* or schizoaffective or psychosis or psychotic or bipolar disorder* or personality disorder*).ti. | 122262 |
| 5 | exp *psychosis/ or exp *acute psychosis/ or exp *chronic psychosis/ or exp *"paranoia (psychosis)"/ or exp *schizophrenia/ or exp *personality disorders/ or exp *bipolar disorder/ | 161261 |
| 6 | or/1-5 | 177601 |
| 7 | (internet or online or digital or web* or social media).ti. | 61281 |
| 8 | exp *Internet/ | 23697 |
| 9 | or/7-8 | 68872 |
| 10 | exp *health information/ | 1783 |
| 11 | exp *information seeking/ or *computer searching/ | 7324 |
| 12 | exp *health literacy/ | 3337 |
| 13 | *health knowledge/ | 6824 |
| 14 | or/10-13 | 18211 |
| 15 | 6 and 9 and 14 | 8 |

##### CINAHL

Host: EbscoHOST

Data parameters: CINAHL 1981 onwards

Date range searched: 1981 to present (Update unknown)

Date searched: 16/06/2022

Searcher: CD

Hits: n=42

| S17 | S6 AND S9 AND S16 | 42 |
| --- | --- | --- |
| S16 | S10 OR S11 OR S12 OR S13 OR S14 OR S15 | 71,217 |
| S15 | (MM "Health Knowledge") | 16,703 |
| S14 | (MM "Health Literacy") | 4,086 |
| S13 | (MM "Health Education") | 15,410 |
| S12 | (MM "Patient Education") | 25,236 |
| S11 | (MM "Information Seeking Behavior") | 2,677 |
| S10 | (MM "Access to Information+") | 10,430 |
| S9 | S7 OR S8 | 115,736 |
| S8 | (MM "Internet+") | 62,333 |
| S7 | TI internet or online or digital or web* or "social media") | 86,218 |
| S6 | S1 OR S2 OR S3 OR S4 OR S5 | 129,725 |
| S5 | (MM "Psychotic Disorders+") OR (MM "Personality Disorders+") | 120,415 |
| S4 | TI schizophren* or schizoaffective or psychosis or psychotic or "bipolar disorder*" or "personality disorder*") | 39,522 |
| S3 | TI SMI | 233 |
| S2 | TI "severe mental illness*" OR "severe mental disorder*" | 1,815 |
| S1 | TI "serious mental illness*" or "serious mental disorder*") | 1,637 |

#### Additional search 2: Peer support for health information

##### MEDLINE

Host: Ovid

Data parameters: Ovid MEDLINE® ALL

Date range searched: 1946 to present (Daily update)

Date searched: 20/06/2022

Searcher: CD

Hits: n=17

| 1 | (serious mental illness* or serious mental disorder*).ti. | 1985 |
| --- | --- | --- |
| 2 | (severe mental illness* or severe mental disorder*).ti. | 2864 |
| 3 | SMi.ti. | 274 |
| 4 | (schizophren* or schizoaffective or psychosis or psychotic or bipolar disorder* or personality disorder*).ti. | 135530 |
| 5 | exp *"bipolar and related disorders"/ or exp *"schizophrenia spectrum and other psychotic disorders"/ or exp *"personality disorders"/ | 187648 |
| 6 | 1 or 2 or 3 or 4 or 5 | 213290 |
| 7 | (peer* adj1 (support* or group* or led)).ti. | 2482 |
| 8 | (expert patient* or "expert* by experience*" or lay expert*).ti. | 166 |
| 9 | exp *peer group/ | 10452 |
| 10 | 7 or 8 or 9 | 11809 |
| 11 | *Access to Information/ | 3801 |
| 12 | *Information Seeking Behavior/ | 1982 |
| 13 | *Patient Education as Topic/ | 40825 |
| 14 | exp *Health Education/ | 152860 |
| 15 | 11 or 12 or 13 or 14 | 157883 |
| 16 | 6 and 10 and 15 | 17 |

##### PsycINFO

Host: Ovid

Data parameters: PsycINFO 1806 to present

Date range searched: 1806 to present (Weekly update)

Date searched: 20/06/2022

Searcher: CD

Hits: n=0

| 1 | (serious mental illness* or serious mental disorder*).ti. | 2052 |
| --- | --- | --- |
| 2 | (severe mental illness* or severe mental disorder*).ti. | 2622 |
| 3 | SMi.ti. | 66 |
| 4 | (schizophren* or schizoaffective or psychosis or psychotic or bipolar disorder* or personality disorder*).ti. | 122262 |
| 5 | exp *"bipolar and related disorders"/ or exp *"schizophrenia spectrum and other psychotic disorders"/ or exp *"personality disorders"/ | 25212 |
| 6 | 1 or 2 or 3 or 4 or 5 | 138879 |
| 7 | (peer* adj1 (support* or group* or led)).ti. | 2562 |
| 8 | (expert patient* or "expert* by experience*" or lay expert*).ti. | 92 |
| 9 | exp *peer relations/ or *peer counseling/ or *peers/ | 22823 |
| 10 | 7 or 8 or 9 | 23745 |
| 11 | exp *health information/ | 1783 |
| 12 | exp *information seeking/ | 5911 |
| 13 | exp *health literacy/ | 3337 |
| 14 | *health knowledge/ | 6824 |
| 15 | 11 or 12 or 13 or 14 | 16848 |
| 16 | 6 and 10 and 15 | 0 |

##### CINAHL

Host: EbscoHOST

Data parameters: CINAHL 1981 onwards

Date range searched: 1981 to present (Update unknown)

Date searched: 20/06/2022

Searcher: CD

Hits: n=4

| S19 | S11 AND S18 | 4 |
| --- | --- | --- |
| S18 | S12 OR S13 OR S14 OR S15 OR S16 OR S17 | 71,233 |
| S17 | (MM "Health Knowledge") | 16,707 |
| S16 | (MM "Health Literacy") | 4,089 |
| S15 | (MM "Health Education") | 15,413 |
| S14 | (MM "Patient Education") | 25,241 |
| S13 | (MM "Information Seeking Behavior") | 2,677 |
| S12 | (MM "Access to Information+") | 10,434 |
| S11 | S6 AND S10 | 203 |
| S10 | S7 OR S8 OR S9 | 8,777 |
| S9 | (MM "Peer Counseling") OR (MM "Peer Group") | 7,678 |
| S8 | TI "expert patient*" OR "expert* by experience* OR "lay expert*") | 137 |
| S7 | TI peer N1 (support* or group* or led) | 2,374 |
| S6 | S1 OR S2 OR S3 OR S4 OR S5 | 144,416 |
| S5 | (MM "Psychotic Disorders+") OR (MM "Personality Disorders+") | 120,459 |
| S4 | TI schizophren* or schizoaffective or psychosis or psychotic or "bipolar disorder*" or "personality disorder*" | 67,417 |
| S3 | TI SMI | 233 |
| S2 | TI "severe mental illness*" OR "severe mental disorder*" | 2,157 |
| S1 | TI "serious mental illness*" or "serious mental disorder*") | 1,779 |
