## Supplemental File 2, and will be used for the link to the file on the preprint slide for "A realist review of medication optimisation of community dwelling service users with serious mental illness"

### Supplementary File 2: Documents included

This table provides full details of the documents included in the review

Table 1: MEDIATE Table of Included Documents

| **Title** | **First author** | **Year** | **Population studied** | **Manuscript Type** | **Summary of findings** |
| --- | --- | --- | --- | --- | --- |
| Shared decision making in mental health: the importance for current clinical practice | Alguera-Lara | 2017 | Serious Mental Illness/Schizophrenia | Narrative review | After culling, 18 relevant articles were included. Themes identified included models of psychiatric care, benefits for patients, and barriers. There is a paucity of published studies specifically related to antipsychotic medications. Shared decision making is a central part of the recovery paradigm and is of increasing importance in mental health service delivery. The field needs to better understand the basis on which decisions are reached regarding psychiatric treatments. Discrete choice experiments might be useful to inform the development of tools to assist shared decision making in psychiatry. |
| A qualitative study of online mental health information seeking behaviour by those with psychosis | Aref-Adib, | 2016 | Psychosis | Qualitative - Semi structured interviews | Results: Mental health related Internet use was widespread. Eighteen people described searching the Internet to help them make sense of their psychotic experiences, and to read more information about their diagnosis, their prescribed psychiatric medication and its side-effects. Whilst some participants sought ‘expert’ online information from mental health clinicians and research journals, others described actively seeking first person perspectives. Eight participants used this information collaboratively with clinicians and spoke of the empowerment and independence the Internet offered them. However nine participants did not discuss their use of online mental health information with their clinicians for a number of reasons, including fear of undermining their clinician’s authority. For some of these people concerns over what they had read led them to discontinue their antipsychotic medication without discussion with their mental health team. |
| Antipsychotic treatment – a systematic literature review and meta-analysis of qualitative studies | Bjornestad | 2020 | People taking antipsychotic medications | Systematic literature review and meta-analysis of qualitative studies | Four meta-themes were identified: (1) short-term benefits; (2) adverse effects and coping processes; (3) surrender and autonomy; (4) long-term compromise of functional recovery. While largely positive about acute and short-term use, patients are more skeptical about using antipsychotic drugs in the longer term. The latter specifically relates to processes of functional and social recovery. The clinical conversations about antipsychotic medication need to include evaluations of contexts of patient experience level, patient autonomy processes, patient values and risk preferences, and patient knowledge and knowledge needs in addition to assessing the severity of symptoms of psychosis. |
| How clients solicit medication changes in psychiatry | Bolden | 2019 | Clients who have been on various psychotropic medications for several years | Qualitative | The analysis shows that clients solicit medication changes at activity boundaries and design them in one of the following ways: reporting a physical problem; reporting a medication problem; explicitly requesting a medication change; and demanding a change. These formats put pressure on the psychiatrist to respond by either offering a solution to the client’s problem or by accepting or rejecting the client’s request. Through a detailed analysis of clients’ communicative behaviours, we show that, in soliciting a medication change, clients ordinarily respect boundaries of medical authority and present themselves as “good” patients who are reliable witnesses of their own experiences. Overall, the paper advances our understanding of patient advocacy in psychiatry and mental health interactions more generally. |
| The quest for well-being: a qualitative study of the experience of taking antipsychotic medication | Carrick | 2004 | Adults taking antipsychotic medication and have a diagnosis of schizophrenia (any subtype), schizoaffective disorder, psychotic illness or borderline personality disorder | Qualitative - Interviews and focus groups | Results indicated that people taking antipsychotic medication do not see side effects and symptoms as separate issues. Instead, they describe drugs as ‘good’ or ‘terrible’—an indication of the total impact of their treatment. The model constructed reflects this, having the core concept of Well-being: that is, normality of function, feeling anappearance to the outside world. Major themes relating to this core category were managing treatment, evaluating treatment and understanding of the situation. |
| A qualitative study of medication adherence amongst people with schizophrenia | Clifford | 2020 | Schizophrenia | Qualitative - Interviews | Consumer-related factors, medication related factors and service-related factors were reported to influence adherence behavior. These included poor insight, unpleasant medication side effects, inadequate efficacy and poor therapeutic alliance. Lessons gained during periods of non-adherence were the motivator for future adherence; such as worsening of symptoms if medication was not taken. Potential implications of future adherence described by Interviewees include greater involvement of peer workers, as they were considered to work more effectively with consumers to encourage adherence. Peer workers had more credibility than other service providers due to their lived experience. Multiple factors were identified that impact on antipsychotic medication adherence, providing opportunities for interventions and improvements in services that would enhance adherence. |
| A feasibility study of expert patient and community mental health team led bipolar psychoeducation groups: implementing an evidence based practice | Coulthard | 2013 | Bipolar | Qualitative | Barriers and enablers were identified at organisational, educational, treatment content, facilitator and patient levels. All barriers under the control of the research team were addressed with subsequent improvements in patient knowledge about the condition and about local service. In addition, self-management, agency and altruism were enhanced. Barriers that could not be addressed required senior clinical and education leadership outside the research team’s control. PS and professional facilitators were successfully trained and worked together to deliver groups which were generally reported as being beneficial. Psychoeducation groups involving CMHT and PS facilitators is acceptable and feasible but their sustainment requires senior leadership within and outside the organisation that control finance and education services. |
| [An analysis of views about supported reduction or discontinuation of antipsychotic treatment among people with schizophrenia and other psychotic disorders.](https://doi.org/10.1186/s12888-022-03822-5) | Crellin | 2022 | Schizophrenia, Schizoaffective disorder, Delusional disorder, Other psychotic disorder | Mixed methods - included semi-structured interviews and collecting quantitative data | We interviewed 269 participants. 33% (95% CI, 27 to 39%) were content with taking long-term antipsychotic medication. Others reported they took it reluctantly (19%), accepted it on a temporary basis (24%) or actively disliked it (18%). 31% (95% CI, 25 to 37%) said they would like to try to stop medication with professional support, and 45% (95% CI, 39 to 51%) wanted the opportunity to reduce medication. People who wanted to discontinue had more negative attitudes towards the medication but were otherwise similar to other participants. Wanting to stop or reduce medication was motivated mainly by adverse effects and health concerns. Professional support was identified as potentially helpful to achieve reduction. This large study reveals that patients are commonly unhappy about the idea of taking antipsychotics on a continuing or life-long basis. Professional support for people who want to try to reduce or stop medication is valued. |
| Facilitators and barriers to the active participation of clients with serious mental illnesses in medication decision making: the perceptions of young adult clients | Delman | 2015 | Young adults with Severe Mental Illness | Qualitative - Interviews | Respondents reported that the primary facilitators to active participation were the psychiatrist’s openness to the client’s perspective, the psychiatrist’s availability outside of office hours, the support of other mental health providers, and personal growth and self-confidence of the young adults. The primary barriers to active participation reported were the resistance of the psychiatrist, the lack of time for consultations, and limited client self-efficacy. Young adults with SMI can be active participants in making decisions about their psychiatric treatment. |
| Shared decision making: People with severe mental illness experiences of involvement in the care of their physical health | Ehrlich | 2017 | lived experience of severe mental illness | Qualitative - Semi-structured face to face interviews | Four key areas in which people with SMI were able to be involved in their health were identified: care continuity within a fragmented care system; medication management; credibility and being mastered; and self-mastery and self-managing health. Shared decision making in mental health care can contribute to equality, control and recovery. Involving people with SMI in shared decision making will contribute positively to their overall health. However, substantial changes are required to shift the health system from a traditional “health professional as expert” approach to one with the patients in the centre. |
| Peer-delivered services and peer support reaching people with schizophrenia: A scoping review | Evans | 2021 | schizophrenia | Scoping review | The 22 studies reviewed described 20 unique interventions, representing significant diversity in formats and components, including those focused on psychological coping and practical skills, such as for independent living. Seven of nine studies measuring at least one key outcome (acute care utilization, patient functioning, positive or negative symptoms) found significant effects favoring peer-delivered intervention. Three of four studies measuring negative symptoms reported a reduction, notable considering these are often difficult to change. Results suggest areas for improvement, including a more thorough examination of program components and tailoring to the specific needs of PWS. |
| Patient subjective experience of treatment with long-acting injectable antipsychotics: a systematic review of qualitative studies | Fiore | 2021 | Patients on long-acting injectable antipsychotics | Systematic review of qualitative studies | Some recurrent issues were associated with LAIs, such as fear of coercion, fear of needles and lack of knowledge about depot therapy. These topics are linked to each other and the patients most concerned about the disadvantages of LAIs are those who are less informed about them, or who have experienced coercion and trauma during hospitalisation. On the other hand, patients who had already received LAIs, and those who had a good therapeutic relationship with their healthcare providers expressed satisfaction with this form of treatment and its continuation. |
| The role of shared decision-making in improving adherence to pharmacological treatments in patients with schizophrenia: a clinical review | Fiorillo | 2020 | Schizophrenia | Critical appraisal of recent literature | SDM is recognized as a promising strategy to improve collaboration between clinicians and patients in achieving recovery. When considering drug treatments, clinicians must evaluate the patient’s preferences, expectations and concerns towards the development of a personalized treatment strategy. Moreover, an active involvement in the decision process could reduce the patient’s perception of being coerced into the use of LAIs. Involving patients in the choice of therapy is not sufficient to increase pharmacological adherence if, at the same time, there is no constant work of comparison and communication with the reference psychiatric team. SDM can be particularly effective for LAI prescription, since patient can have prejudices and unjustified fears related to the LAI formulation, which the doctor must resolve. |
| Personal Accounts of Discontinuing Neuroleptic Medication for Psychosis | Geyt | 2017 | Patients who had taken neuroleptic medication for psychosis for at least 3 months | Qualitative - semi-structured interviews | We used a grounded theory approach to analyze transcripts from interviews with 12 participants. We present a preliminary grounded theory of the processes involved in making choices about neuroleptic medication. We identified three tasks as important in mediating participants’ choices: (a) forming a personal theory of the need for, and acceptability of taking, neuroleptic medication; (b) negotiating the challenges of forming alliances with others; and (c) weaving a safety net to safeguard well-being. Progress in the tasks reflected a developmental trajectory of becoming an expert over time and was influenced by systemic factors. Our findings highlight the importance of developing resources for staff to facilitate service user choice. |
| Understanding how clinician-patient relationships and relational continuity of care affect recovery from serious mental illness: STARS study results | Green | 2008 | Schizophrenia, Schizoaffective disorder, Affective psychosis, Bipolar | Qualitative - mixed methods, exploratory, longitudinal study - questionnaires and interviews | Qualitative data showed that positive, trusting relationships with clinicians, developed over time, aid recovery. When “fit” with clinicians was good, long-term relational continuity of care allowed development of close, collaborative relationships, fostered good illness and medication management, and supported patient-directed decisions. Most valued were competent, caring, trustworthy, and trusting clinicians who treated clinical encounters “like friendships,” increasing willingness to seek help and continue care when treatments were not effective and supporting “normal” rather than “mentally ill” identities. Statistical models showed positive relationships between recovery-oriented patient-driven care and satisfaction with clinicians, medication satisfaction, and recovery. Relational continuity indirectly affected quality of life via satisfaction with clinicians; medication satisfaction was associated with fewer symptoms; fewer symptoms were associated with recovery and better quality of life. |
| The role of trust and hope in antipsychotic medication reviews between GPs and service users: a realist review | Grunwald | 2021 | Schizophrenia/Psychosis | Realist review | Meaningful Antipsychotic medication reviews may not occur for individuals with only primary care medical input. Several, often mutually reinforcing, mechanisms have been identified as potential barriers to conducting such reviews, including low expectations of recovery for people with severe mental illness, a perceived lack of capability to understand and participate in medication reviews, linked with a lack of information shared in appointments between GPs and Service Users, perceived risk and uncertainty regarding antipsychotic medication and illness trajectory. The review identified reciprocal and reinforcing stereotypes affecting both GPs and service users. Possible mechanisms to counteract these barriers are discussed, including realistic expectations of medication, and the need for increased information sharing and trust between GPs and service users. |
| Re-starting the conversation: improving shared decision making in antipsychotic prescribing | Grunwald and Thompson | 2021 | Schizophrenia/Psychosis | Commentary | Shared Decision-Making (SDM) is one of the key components of patient-centred care. People diagnosed with schizophrenia/psychosis still face significant barriers to achieving this, particularly when it comes to antipsychotic medication prescribing. These barriers include issues such as stigma, feelings of coercion and lack of information. Clinicians also describe barriers to achieving SDM in antipsychotic prescribing, including a lack of training and support. In this viewpoint article, we provide a summary of these barriers from the perspectives of both service users and clinicians based. We suggest that, to make a practical first step towards achieving SDM, the conversation around antipsychotic prescribing needs to be re-started. However, the onus to do this should not be placed solely on the shoulders of Service Users. More research is needed to address this issue. |
| Nonadherence with antipsychotic medication in schizophrenia: challenges and management strategies | Haddad | 2014 | Schizophrenia | Commentary | Nonadherence lies on a spectrum, is often covert, and is underestimated by clinicians, but affects more than one third of patients with schizophrenia per annum. It increases the risk of relapse, rehospitalization, and self-harm, increases inpatient costs, and lowers quality of life. It results from multiple patient, clinician, illness, medication, and service factors, but a useful distinction is between intentional and unintentional nonadherence. There is no gold standard approach to the measurement of adherence as all methods have pros and cons. Interventions to improve adherence include psychoeducation and other psychosocial interventions, antipsychotic long-acting injections, electronic reminders, service-based interventions, and financial incentives. These overlap, all have some evidence of effectiveness, and the intervention adopted should be tailored to the individual. Psychosocial interventions that utilize combined approaches seem more effective than unidimensional approaches. There is increasing interest in electronic reminders and monitoring systems to enhance adherence, eg, Short Message Service text messaging and real-time medication monitoring linked to smart pill containers or an electronic ingestible event marker. Financial incentives to enhance antipsychotic adherence raise ethical issues, and their place in practice remains unclear. Simple pragmatic strategies to improve medication adherence include shared decision-making, regular assessment of adherence, simplification of the medication regimen, ensuring that treatment is effective and that side effects are managed, and promoting a positive therapeutic alliance and good communication between the clinician and patient. These elements remain essential for all patients, not least for the small minority where vulnerability and risk issue dictate that compulsory treatment is necessary to ensure adherence. |
| How Do People Experience Early Intervention Services for Psychosis? A Meta-Synthesis | Hansen | 2018 | Psychosis | Meta-synthesis | A broad literature search was performed in June and July 2016. After screening, 17 qualitative studies were included. We analyzed the findings in two main steps: (a) translating studies into one another and (b) synthesizing the findings from the studies. Through these interpretative processes, we found five new and overarching themes: (a) something is wrong, (b) do for myself, (c) it’s about people, (d) a price to pay, and (e) ongoing vulnerability. We describe these themes as a process that service users’ maneuver through in their contact with the services. Our findings are discussed in light of relevant research. |
| Mental health professionals experiences with shared decision making for patients with psychotic disorders: a qualitative study | Haugom | 2020 | Health Professionals who work at mental health centres where patients with psychotic disorders are treated | Qualitative - focus group interviews | Health professionals primarily understand the SDM concept to mean giving patients information and presenting them with a choice between different antipsychotic medications. Among the barriers to SDM, they emphasized that patients with psychosis have a limited understanding of their health situation and that time is needed to build trust and alliances. Health professionals mainly understand patients with psychotic disorders as a group with limited abilities to make their own decisions. They also described the concept of SDM with little consideration of presenting different treatment options. Psychological or social interventions were often presented as complementary to antipsychotic medications, rather than as an alternative to them. |
| Long-acting injectable (depot) antipsychotics and changing treatment philosophy: Possible contribution to integrative care and personal recovery of schizophrenia | Jakovljevic | 2014 | Schizophrenia | Commentary | Despite a growing consensus based on number naturalistic studies for the superiority of LAAIs over oral equivalents due to improved adherence and more stable pharmacokinetics, many current official treatment guidelines recommend basic treatment with oral antipsychotics. In recent years the interest of using SGLAAIs, particularly in the early stages of schizophrenia, has increased in order to improve treatment outcome and prognosis of illness. Schizophrenia should be treated with antipsychotics earlier and for longer periods of time, and thus LAIAs may be optimal choice what is also suggested in some recent treatment guidelines. Any patient for whom long-term antipsychotic treatment is indicated should be offered LAAIs. Given the well recognised relationship between non-adherence and risk of relapse, patients who are irregular in taking medications are must-offer candidates for LAAIs. |
| Medication nonadherence in bipolar disorder: a narrative review | Jawad | 2018 | Bipolar Disorder | Narrative review | Factors associated with nonadherence include adverse effects of medication, complex medication regimens, negative patient attitudes to medication, poor insight, rapid-cycling BD, comorbid substance misuse and a poor therapeutic alliance. Clinicians should routinely enquire about nonadherence in a non-judgmental fashion. Potential steps to improve adherence include simple pragmatic strategies related to prescribing including shared decision making, psychoeducation with a clear focus on adherence, reminders (traditional and digital), potentially using a depot rather than an oral antipsychotic, managing comorbid substance misuse and improving therapeutic alliance. Financial incentives have been shown to improve adherence to depot antipsychotics, but this approach raises ethical issues and its long-term effectiveness is unknown. Often a combination of approaches will be required. The strategies that are adopted need to be patient specific, reflecting that nonadherence has no single cause, and chosen by the patient and clinician working together. |
| Making decisions about antipsychotics: a qualitative study of patient experience and the development of a decision aid | Kaar | 2019 | Chronic psychotic illness | Qualitative - focus groups | Twenty-three patients participated in the study. Thematic analysis revealed that ‘adverse effects’ was the most common theme identified by patients surrounding antipsychotic medication decision-making followed by ‘mode and time of administration’, ‘symptom control’ and ‘autonomy’. The final decision aid is included to provoke further discussion and development of such aids. Patients commonly report negative experiences of antipsychotic medication, in particular side-effects, which remain critical to future decision making around antipsychotic medication. Clinical encounters that increase patient knowledge and maximise autonomy in order to prevent early negative experiences with antipsychotic medication are likely to be beneficial. |
| Barriers and enablers to shared decision making in psychiatric medication management: A qualitative investigation of clinician and service users’ views. | Kaminskiy | 2021 | Severe mental illness | Qualitative - semi-structured interviews | The results offer a detailed contextualized account of how medication decisions are made. For psychiatrists and service user participants SDM is seen as a way of enhancing service users’ engagement in and control over treatment decisions. While psychiatrists value the transactional benefits of SDM, service user participants and psychiatric nurses conceptualize SDM as a long-term endeavor embedded within therapeutic partnerships. For service users these partnerships mitigate acknowledged problems of feeling unable to be fully involved during times of crisis. This study identified a range of barriers and facilitators to SDM concerning psychiatric medications from the lived experience of service users and the professional experience of clinicians. Furthermore, it indicates new potential intervention points to support SDM in psychiatric medication decisions. |
| Retrospective Accounts of the Process of Using and Discontinuing Psychiatric Medication | Katz | 2019 | Serious mental illness | Qualitative - semi-structured interviews | The study was carried out using the narrative approach to life stories method. Participants were 12 women and 9 men who had discontinued their prescribed medication following psychiatric hospitalization. Four main themes were revealed in the data analysis: (a) the experience with medication, (b) the process of discontinuing medication, (c) elements that helped achieve successful medication discontinuation, and (d) the perceived impact of medication discontinuation. Our findings challenge the widespread notion that discontinuing psychiatric medication is necessarily negative and suggest that, for some, it is a legitimate and meaningful life choice. |
| Mental health service users experiences of medication discontinuation: a systematic review of qualitative studies | Keogh | 2022 | People with severe and enduring mental health difficulties on common long term medications such as antipsychotics, mood stabilisers and antidepressants | Systematic review of qualitative studies | Six themes were identified: (1) Taking medications: a loss of autonomy, (2) Discontinuing medication: a thought-out process, (3) Factors influencing the decision to discontinue medication, (4) Discontinuing medication: experiences of the process, (5) Outcomes of discontinuing medication, (6) Managing mental distress in the absence of medication. Service providers need to be aware that for some service user’s psychotropic medication is not deemed a suitable treatment approach. Those who wish to discontinue medication need to be supported in the context of positive, therapeutic risk where their mental and physical health can be monitored and the likelihood of success increased. |
| “I found hundreds of other people…but I still wasn’t believed” – An exploratory study on lived experiences of antipsychotic withdrawal | King | 2022 | Prescibed and taken antipsychotic drug(s) for any indication | Qualitative - semi-structured interviews | The themes were: balancing priorities, withdrawal journey, invalidation of experiences and peer community. Findings show the extent of symptoms that can be experienced secondary to antipsychotic discontinuation, they highlight the wider individual context that antipsychotics impact, and the meaningful reasoning behind withdrawal decisions. |
| Service-user efforts to maintain their wellbeing during and after successful withdrawal from antipsychotic medication | Larsen-Barr | 2021 | People who had discontinued antipsychotics - sample obtained included bipolar disorder, depression, ocd | Qualitative - semi-structured interviews | Of the seven women who volunteered to participate, six reported bipolar disorder diagnoses and one reported diagnoses of obsessive compulsive disorder and depression. The women reported successfully discontinuing antipsychotics for 1.25–25 years; six followed a gradual withdrawal method and had support to prepare for and manage this. Participants defined wellbeing in terms of their ability to manage the impact of any difficulties faced rather than their ability to prevent them entirely, and saw this as something that evolved over time. They described managing the process and maintaining their wellbeing afterwards by ‘understanding myself and my needs’, ‘finding what works for me’ and ‘connecting with support’. Sub-themes expand on the way in which they did this. For example, ‘finding what works for me’ included using a tool-box of strategies to flexibly meet their needs, practicing acceptance, drawing on persistence and curiosity and creating positive life experiences. |
| Attempting to stop antipsychotic medication: success, supports and efforts to cope | Larsen-Barr | 2018 | Adults who were taking or had taken antipsychotic medication for at least 3 months for any reason | Survey | Among the 105 people who had attempted discontinuation, 61.9% described unwanted withdrawal effects and 27.6% of the group described psychotic or manic relapse during the withdrawal period. Within this group 55% described successfully stopping all AM for varying lengths of time, half reported no current use, and half described having some form of professional, family, friend, and/or service-user or peer support for their attempt. Having support was positively associated with success and negatively associated with both current use, and relapse during withdrawal. A range of coping efforts were described, but having coping strategies failed to show significant associations with any of the dependent variables explored. Among those who described successfully stopping, some described returning to AM for short periods when needed while others reported managing well with alternative methods alone. |
| Determinants of adherence to treatment in first-episode psychosis: a comprehensive review | Leclerc | 2015 | First episode psychosis (FEP) | A comprehensive review | FEP = First Episode Psychosis  A total of 157 articles were screened, of which 33 articles were retained for full review. The factors related to nonadherence were: a) patient-related (e.g., lower education level, persistent substance use, forensic history, unemployment, history of physical abuse); b) environment-related (e.g., no family involved in treatment, social adjustment difficulties); c) medication-related (e.g., rapid remission of negative symptoms when starting treatment, therapeutic alliance); and d) illness-related (e.g., more positive symptoms, more relapses). Treatment factors that improve adherence include a good therapeutic alliance and a voluntary first admission when hospitalization occurs. The results of this review suggest that nonadherence to treatment in FEP is multifactorial. Many of these factors are modifiable and can be specifically targeted in early intervention programs. Very few studies have assessed strategies to raise adherence in FEP. |
| A qualitative exploration of family members’ perspectives on reducing and discontinuing antipsychotic medication | Lewins | 2022 | Family members of people with psychosis | Qualitative - semi-structured interviews | The majority of family members valued antipsychotic medication primarily in supporting what they saw as a fragile stability in the person they cared for. Their views of medication were ambivalent, combining concerns about adverse effects with a belief in the importance of medication due to fears of relapse. They described a need for constant vigilance in relation to medication to ensure it was taken consistently, and often found changes, particularly reduction in medication difficult to contemplate. Findings highlight that family members’ attitudes to medication sometimes conflict with those of the people they care for, impacting on their health and the caring relationship. Family members may need more support and could be usefully involved in medication decision-making. |
| The Effect of Therapeutic Alliance on Attitudes Toward Psychiatric Medications in Schizophrenia | Lim | 2021 | Schizophrenia and Schizoaffective disorder | Quantitative | Older age, longer duration of illness, presence of medical comorbidities, lower levels of internalized stigma, higher levels of insight, higher levels of functioning, lesser severity of depressive symptoms, and positive symptoms were found to be significantly associated with greater levels of drug attitude (small to moderate associations). Only therapeutic alliance had a large correlation with drug attitude (ρ = 0.503, P < 0.001). The therapeutic alliance scores between the 2 health care professionals groups are not significantly different. However, participants who have identified psychiatrists as the health care professional that contributed the most to their recovery reported a significantly more positive attitude (μ = 6.18, SD = 3.42) toward psychiatric medication as compared with the other health care professionals group (μ = 3.11, SD = 5.32, P = 0.004). Only 2 factors, the Revised Helping Alliance Questionnaire (β = 0.424, P < 0.001) and Personal and Social Performance scale (β = 0.272, P = 0.006), were statistically significant predictors of drug attitude. |
| Adherence to antipsychotic medication in bipolar disorder and schizophrenic patients: A systematic review | Garcia | 2016 | Schizophrenia, Bipolar Disorder | Systematic review | Analyzing 38 studies conducted in a total of 51,796 patients, including patients with schizophrenia spectrum disorders and bipolar disorder, we found that younger age, substance abuse, poor insight, cognitive impairments, low level of education, minority ethnicity, poor therapeutic alliance, experience of barriers to care, high intensity of delusional symptoms and suspiciousness, and low socioeconomic status are the main risk factors for medication nonadherence in both types of disorder. In the future, prospective studies should be conducted on the use of personalized patient tailored treatments, taking into account risk factors that may affect each individual, to assess the ability of such approaches to improve adherence and hence prognosis in these patients. |
| Effective Strategies for Nurses Empowering Clients With Schizophrenia: Medication Use as a Tool in Recovery | Mahone | 2016 | Schizophrenia | Commentary | Clients with schizophrenia require maintenance treatment with antipsychotic medication and psychosocial therapy to maintain symptom control. Rates of medication adherence or follow-through are low in clients with schizophrenia. This increases the risk of relapse and contributes to poor quality of life. As educators and advisers, psychiatric nurses can collaborate with clients to improve adherence and other outcomes using shared decision-making techniques and tools that engage and empower clients to actively participate in decisions about their treatment. This article outlines effective strategies used by psychiatric nurses to improve outcomes in clients with schizophrenia and uses a case example for demonstrating this strategy in a client with schizophrenia. |
| The clinical characterization of the patient with primary psychosis aimed at personalization of management | Maj | 2021 | Primary Psychosis | Commentary | Although many mental health services would declare themselves “recovery-oriented”, it is not common that a focus on empowerment, identity, meaning and resilience is ensured in ordinary practice. The present paper aims to address this situation. It describes systematically the salient domains that should be considered in the characterization of the individual patient with primary psychosis aimed at personalization of management. These include positive and negative symptom dimensions, other psychopathological components, onset and course, neurocognition and social cognition, neurodevelopmental indicators; social functioning, quality of life and unmet needs; clinical staging, antecedent and concomitant psychiatric conditions, physical comorbidities, family history, history of obstetric complications, early and recent environmental exposures, protective factors and resilience, and internalized stigma. For each domain, simple assessment instruments are identified that could be considered for use in clinical practice and included in standardized decision tools. A management of primary psychosis is encouraged which takes into account all the available treatment modalities whose efficacy is supported by research evidence, selects and modulates them in the individual patient on the basis of the clinical characterization, addresses the patient’s needs in terms of employment, housing, self-care, social relationships and education, and offers a focus on identity, meaning and resilience. |
| The collaborative management of antipsychotic medication and its obstacles: A qualitative study | Martinez-Hernaez | 2020 | Patients (users of antipsychotics), family caregivers and mental health professionals | Qualitative - semi-structured interviews and focus groups | We detected three main obstacles to collaboration among participants. First, different understanding of the patient's distress, either as deriving from the symptoms of the disorder (professionals) or the adverse effects of the medication (patients). Second, differences in the definition of (un)awareness of the disorder. Whereas professionals associated disorder awareness with treatment compliance, caregivers understood it as synonymous with self-care, and among patients “awareness of suffering” emerged as a comprehensive category of a set of discomforts (i.e., symptoms, adverse effects of medication, previous admissions, stigma). Third, discordant expectations regarding clinical communication that can be condensed in the differences in meaning between the Spanish words “trato” and “tratamiento”, where the first denotes having a pleasant manner and agreement, and the second handling and management. We conclude that these three obstacles pave the way for coercive practices and promote patients' de-subjectivation, named here as the “total patient” effect. |
| Decision making in recovery-oriented mental health care | Matthias | 2012 | Providers - Psychiatrists and nurse   Consumers - People with severe mental illness | Qualitative | Providers initiated most decisions, although they often invited consumers to participate in decision making. Decisions initiated by consumers elicited a greater degree of discussion and disagreement, but also frequently resulted in consumers’ preferences prevailing. Consultations generally exhibited more characteristics of person-centeredness than SDM. |
| The least worst option: user experiences of antipsychotic medication and lack of involvement in medication decisions in a UK community sample | Morant | 2018 | People with a psychotic condition who are taking or have taken antipsychotics within last three months | Qualitative - semi-structured interviews | Antipsychotic medication was perceived to have beneficial effects on symptoms and relapse risk, but adverse effects were prominent, including a global state of lethargy and demotivation. Weighing these up, the majority viewed antipsychotics as the least worst option. Participants were split between positions of ‘‘willing acceptance’’, ‘‘resigned acceptance’’ and ‘‘non-acceptance’’ of taking antipsychotics. Many felt their choices about medication were limited, due to the nature of their illness or pressure from other people. They commonly experienced their prescribing psychiatrist as not sufficiently acknowledging the negative impacts of medication on life quality and physical health concerns and described feeling powerless to influence decisions about their medication. |
| Rethinking medication prescribing practices in an inner-city Hispanic mental health clinic | Opler | 2004 | Psychotic disorders | Unspecified | Improved compliance with pharmacotherapy was achieved in treating Hispanic outpatients with psychotic disorders when recognition of culturally based differences between patients and psychiatrists led to modifications in prescribing practices. Unacculturated Hispanic outpatients experienced akathisia as an increase in “nerviosismo.” Addressing this issue, as well as using anxiolytics and low doses of antipsychotics when beginning treatment, led to an improvement in compliance. Increased discussion of other antipsychotic side effects, which forced us to confront our false assumption that unacculturated Hispanics would be prone to suggestibility and, therefore, that discussions of side effects would lead to an increase in somatization, similarly improved medication compliance and therapeutic alliance. Practicing psychiatrists need to become aware of cultural factors to better treat patients with different backgrounds. |
| An exploratory analysis of the role of social supports in psychiatric medication discontinuation: results related to family involvement | Ostrow | 2019 | Schizophrenia, Schizoaffective disorder, Schizophreniform disorder, Psychosis, Bipolar, Major depressive disorder | Survey Design | Of all social support groups, only family was significantly associated with medication discontinuation. Respondents who rated family as helpful in the discontinuation process were less likely to completely discontinue than those who rated family as unhelpful or who reported no family involvement. Additionally, we observed a statistically significant but nonlinear relationship where respondents who rated their families as either “very supportive” or “very unsupportive” of the decision to discontinue were less likely to meet their original discontinuation goal than those with more neutral ratings. |
| Qualitative systematic review of barriers and facilitators to patient-involved antipsychotic prescribing | Pedley | 2018 | Patients with Serious mental illness  Healthcare professionals who care for at least one person who has serious mental illness | Systematic thematic synthesis | Synthesis of 29 studies identified the following key influences on involvement: patient’s capability, desire and expectation for involvement, organisational context, and the consultation setting and processes. Optimal patient involvement in antipsychotic decisions demands that individual and contextual barriers are addressed. There was divergence in perceived barriers to involvement identified by patients and prescribers. For example, patients felt that lack of time in consultations was a barrier to involvement, something seldom raised by prescribers, who identified organisational barriers. Patients must understand their rights to involvement and the value of their expertise. Organisational initiatives should mandate prescriber responsibility to overcome barriers to involvement. |
| Medication adherence in patients with schizophrenia | Phan | 2016 | Schizophrenia | Commentary | There appears to be no single strategy to manage and prevent medication partial adherence or nonadherence. Based on a review of the literature, interventions that appear to be most effective are multidimensional and address different factors that are specific to the patient. With patient consent, they should involve the patient’s support system, such as caregivers, family, and friends. The barriers to medication adherence will be different for each patient and may change over time. Thus, it is important for providers to acknowledge that patients will, from time to time, become partially adherent or nonadherent. The risk for nonadherence should continually be assessed and addressed to minimize the risk of rehospitalization, relapse, loss of function and an overall worse prognosis. |
| The modern perspective for long-acting injectables antipsychotics in the patient-centered care of schizophrenia | Pietrini | 2019 | Schizophrenia | review (type unspecified) using panel of experts | In the present study, a panel of experts (psychiatrists, psychologists, nurse, and social worker) gathered to review and explore the need for contemporary use of second generation antipsychotic long-acting injectables (SGA LAIs) in “recovery-oriented” and “patient-centered” care of schizophrenia. Starting from the available data and from sharing personal attitudes and experiences, the panel selected three clinical dimensions considered useful in characterizing each patient: phase of disease, adherence to treatment, and level of functioning. For each clinical dimension, perspectives of patients and caregivers with regard to needs, expectations, and personal experiences were reviewed and the role of SGA LAIs in achieving shared goals examined. The experts concluded that from today’s modern perspectives, SGA-LAIs may play an important role in breaking the spiral of desocialization and functional decline in schizophrenia, thus favoring the recovery process. |
| Anti-psychotic medication decision making during pregnancy: A co-produced research study | Pinfold | 2019 | Schizophrenia, Bipolar Disorder | Qualitative - semi-structured interviews | The accounts highlighted decisional uncertainty, with medication decisions situated among multiple sources of influence from self and others. Women retained strong feelings of personal ownership for their decisions, whilst also seeking out clinical opinion and accepting they had constrained choices. Two styles of decision making emerged: shared and independent. Shared decision making involved open discussion, active permission seeking, negotiation and coercion. Independent women-led decision making was not always congruent with medical opinion, increasing pressure on women and impacting pregnancy experiences. A common sense self-regulation model explaining management of health threats resonated with women’s accounts. |
| Communication about adherence to long-term antipsychotic prescribing: an observational study of psychiatric practice | Quirk | 2013 | Patients prescribed antipsychotic medication | Qualitative  Conversation analysis | In 22 (24 %) consultations, partial/non-adherence was disclosed. Most commonly, it was volunteered without prompting and was more likely to be presented as a deliberate choice than omission by the patient. Psychiatrists responded to all but one disclosure, and patients delivered their reports in ways that minimised the prospect of this response being disciplinary. The most common outcome was a change in prescribing: a medication omission, swap or dosage reduction. |
| A thematic analysis assessing clinical decision-making in antipsychotic prescribing for schizophrenia | Roberts | 2018 | Clinicians with experience in prescribing for schizophrenia | Qualitative - semi-structured interviews | The analysis identified five themes underpinning prescribing behaviour: (1) ownership and collaboration; (2) compromise; (3) patient involvement; (4) integrating research evidence; and (5) experience. The themes mapped to various degrees onto current models of evidence-based decision making and suggest that there is scope to re-think the guideline implementation frameworks to incorporate recurring themes salient to clinicians who ultimately use the guidelines. This will further translation of future evidence into clinical practice, accelerating clinical progress. |
| Mediating role of illness representation among social support, therapeutic alliance, experience of medication side effects, and medication adherence in persons with schizophrenia | Rungruangsiripan | 2011 | Schizophrenia | Cross-sectional research study  Interviews and quantitative analysis | Results indicated that therapeutic alliance and the experience of medication side effects enhanced illness representation, which in turn led to an intention to change adherence behavior. Social support did not alter illness representation or adherence behavior. Because illness representation positively influenced patients’ intention to change adherence behavior, mental health nurses should promote patients’ perception about their illness to enhance medication adherence. |
| A narrative meta-synthesis of how people with schizophrenia experience facilitators and barriers in using antipsychotic medication: Implications for healthcare professionals | Salzmann-Erikson | 2018 | schizophrenia or schizophrenia-like conditions | Narrative meta-synthesis | The findings showed that patients were uninformed about medication but valued talks about medication with professionals. The findings also demonstrated that patients are motivated to take medication in order to gain stability in their life and to be able to participate in life activities and in relationships. Good support, both from relatives and professionals, also motivates them to continue taking medication. The obstacles were side effects, pressure and compulsion, and rigid organizations. |
| Sharing decisions in consultations involving anti-psychotic medication: a qualitative study of psychiatrists' experiences | Seale | 2006 | Consultant Psychiatrists | Qualitative - Interviews | This qualitative study reports the views of 21 general adult psychiatrists working in UK about their experiences of consultations involving discussion of antipsychotic medication. Interviewees reported a general commitment to achieving concordant relationships with patients and described a number of strategies they used to promote this. In this respect, their self-perception differs from the picture of authoritarian practice painted by critics of psychiatry, and by some studies reporting patients’ views. Interviewees also described obstacles to achieving concordance, including adverse judgements of patients’ competence and honesty about their medication use. Explaining the adverse effects of medication was perceived to discourage some patients from accepting this treatment. Moments of strategic dishonesty were reported. Psychiatrists perceived that trust could be damaged by episodes of coercion, or by patients’ perception of coercive powers. We conclude that a self-perception of patient-centredness may not preclude psychiatrists from fulfilling a social control function. |
| Cross-sector user and provider perceptions on experiences of shared-care clozapine: a qualitative study | Sowerby | 2017 | Clozapine service users or healthcare professionals caring for clozapine service users | Qualitative - semi-structured interviews and focus groups | CSU = Clozapine Service User HCP = Healthcare Professional  32 HCPs and 6 CSUs were recruited and 14 interviews and 6 participant homogenous focus groups were run. Four shared superordinate themes were identified: Clozapine Process, The Sharing of Care, The Provision of Care and Multi-professional Relationships. Differences between Adult and Forensic engagement in shared care were noted and both HCP and CSU relationships were mapped to the Wish conceptual framework of relationships to provide insight into how shared-care clozapine can provide a mechanism for provision of person-centred care, which was present in the Forensic HCP–CSU but not General Adult HCP–CSU relationship. The Forensic HCP/CSU relationship demonstrated how cross-sector working through shared care clozapine can provide a mechanism for provision of person-centred care by enabling a person-centred focus to care delivery which supported CSUs to live as independently as possible. Person-centred care demonstrably improves patient care outcomes and wider implementation of shared care clozapine could provide greater integration of people with serious mental illness and reduce stigma within the community while improving patient outcomes. |
| Five year outcomes of tapering antipsychotics drug doses in a community mental health centre | Steingard | 2018 | Schizophrenia spectrum disorders, mood disorders and borderline personality disorder | Naturalistic study | Over a period of 6 months, the author invited patients who were clinically stable and able to participate in discussions of potential risks and benefits to begin gradual dose reductions. Initially, 40 expressed interest in tapering and 27 declined. The groups did not differ in age, sex, race, or diagnosis. The group who chose to taper began on significantly lower doses. Most patients succeeded at making modest dose reductions. At 5 years, there were no significant differences in the two outcomes measures, rate of hospitalization and employment status. Many patients were able to engage in these discussions which did not result in widespread discontinuation of drug. This is a naturalistic, small study of a topic that warrants further research. |
| experiences of taking neuroleptic medication and impacts on symptoms sense of self and agency: A systematic review and thematic analysis of qualitative Data | Thompson | 2020 | Schizophrenia, Psychotic disorder, Bipolar disorder and Borderline Personality disorder | Systematic review and thematic synthesis of qualitative data | Neuroleptics were commonly experienced as producing a distinctive state of lethargy, cognitive slowing, emotional blunting and reduced motivation, which impaired functioning but also had beneficial effects on symptoms of psychosis and some other symptoms (e.g. insomnia). For some people, symptom reduction helped restore a sense of normality and autonomy, but others experienced a loss of important aspects of their personality. Across studies, many people adopted a passive stance towards long-term medication, expressing a sense of resignation, endurance or loss of autonomy. |
| Why do psychiatric patients stop antipsychotic medication? A systematic review of reasons for nonadherence to medication in patients with serious mental illness | Velligan | 2017 | Major depressive disorder, Schizophrenia, Bipolar disorder | Systematic review | A qualitative analysis of data from 36 articles identified 11 categories of reasons for nonadherence. Poor insight was identified as a reason for nonadherence in 55.6% (20/36) of studies, followed by substance abuse (36.1%, 13/36), a negative attitude toward medication (30.5%, 11/36), medication side effects (27.8%, 10/36), and cognitive impairments (13.4%, 7/36). A key reason directly associated with intentional nonadherence was a negative attitude toward medication, a mediator of effects of insight and therapeutic alliance. Substance abuse was the only reason consistently associated with unintentional nonadherence, regardless of type and stage of SMI. |
| Deciding to discontinue prescribed psychotropic medication: A qualitative study of service users’ experiences. | Watts | 2021 | Schizophrenia, schizoaffective disorder, bipolar, psychotic disorder | Qualitative - semi-structured interviews | Findings suggest that while medication was useful for many in the short-term, the adverse effects had significant impact and contributed to the decision to come off medication. Participants also reported being driven by a questioning of the biomedical model of treatment and the belief that there were other strategies to manage their distress. Mixed experiences of support from healthcare professionals for the medication cessation process were reported. The discontinuation process was often difficult resulting in changes in mood and behaviour which for many culminated in relapse of distress, rehospitalization and return to medication. To support the process of coming off and staying off medication, participants identified a range of useful strategies but particularly highlighted the importance of peer support. Findings from this study demonstrate the importance of mental health nurses having a collaborative discussion with service users which may support safer decision making and lessen the risk of people discontinuing medication abruptly. Finding also indicates a need for robust studies that develop and test interventions to support people who wish to discontinue psychotropic medications. |
| Redefining Medication Adherence in the Treatment of Schizophrenia: How Current Approaches to Adherence Lead to Misinformation and Threaten Therapeutic Relationships | Weiden | 2016 | Schizophrenia | Review (type unspecified) | This review suggests that the primary goal is to reduce medication gaps and discontinuities as much as possible regardless of cause. Also, even when these gaps happen, it is possible to minimize collateral damage from misinformation by focusing on better detection and disclosure. For patients who are ambivalent or opposed to antipsychotic medication, our belief is that it is best to take a longer term, developmental perspective. The road to better adherence may take a while, and in the long-run it is more effective to focus on the therapeutic relationship. This can be accomplished through dropping adversarial or imperious responses to patients’ concerns, and focus on understanding those concerns before responding. Finally, when nonadherence is a choice and seems inevitable, then it is best to stop trying to “stop” nonadherence, and rather to focus on coming up with a collaborative harm-reduction strategy to try to mitigate the risks and consequences of medication discontinuation. |
| Experiences of antipsychotic use in patients with early psychosis: a two-year follow-up study | Yeisen | 2017 | Psychosis | Qualitative - semi-structured interviews | The textual analysis revealed four main themes that affected adherence largely: 1) Positive experiences of admission, 2) Sufficient timely information, 3) Shared decision-making and 4) Changed attitudes to antipsychotics due to their beneficial effects and improved insight into illness. Patients reported several factors to have a prominent impact on adherence to their antipsychotics. The patients do not independently choose to jeopardize their medication regime. Health care staff play an important role in maintaining good adherence by being empathetic and supportive in the admission phase, giving tailored information according to patients’ condition and involving patients when making treatment decisions. |
| Mental health pharmacists views on shared decision-making for antipsychotics in serious mental illness | Younas | 2016 | Secondary care mental health Pharmacists | Qualitative - semi-structured interviews | Thirteen mental health pharmacists were interviewed. SDM was perceived to be linked to positive clinical outcomes including adherence, service user satisfaction and improved therapeutic relations. Despite more prescribers and service users supporting SDM, it was not seen as being practised as widely as it could be; this was attributed to a number of barriers, most predominantly issues surrounding service user’s lacking capacity to engage in SDM and time pressures on clinical staff. The need for greater effort to work around the issues, engage service users and adopt a more inter-professional approach was conveyed. |
| Factors associated with complete discontinuation of medication among patients with schizophrenia in the year after hospital discharge | Zhou | 2017 | Schizophrenia | Prospective non-interventional observational study | Altogether 25.8% of the sample discontinued medication in the one-year after discharge. Logistic regression analysis showed that shorter duration of illness, lack of health insurance, and poor insight at the time of discharge were significantly associated with complete discontinuation of medication (p < 0.05). Patients discontinued their medication within a year after psychiatric hospitalization which was associated with a lack of insurance coverage, less insight into their illness and shorter duration of illness. Interventions that strengthen patient engagement in treatment through insurance coverage and insight, fostered through psychoeducational intervention, may increase medication compliance. |
| You say “schizophrenia” and I say “psychosis”: just tell me when I can come off this medication | Zipursky | 2020 | Schizophrenia | Literature review | This paper reviews the literature on relapse rates following a first episode of schizophrenia and identifies factors that contribute to the discrepancies in the rates reported. These factors include sampling considerations, the distribution of psychiatric diagnoses, the duration of follow-up, the rate of medication discontinuation and the criteria used to define illness recurrence. We propose that individuals for whom the diagnosis of their first psychotic episode is determined with ongoing follow-up to be due to schizophrenia are at extremely high risk of relapse and should be advised to continue antipsychotic medication for the long-term. Those whose first episode of psychosis is determined to be due to other causes are also at high risk of illness recurrence off medications. Recommendations for maintenance treatment should be tailored to reflect the risk of relapse and sequelae of relapse associated with specific causes of first episode psychosis. |
| Shared risk-taking: shared decision making in serious mental illness. | Zisman-Ilani | 2021 | Serious Mental Illness | Open Forum | This Open Forum presents a reconceptualization of SDM as a process of shared risk taking that often occurs during different phases of illness management and recovery. The concepts of intersubjectivity, meaning making, and metacognition are offered to inform clinical interventions needed to address risk in SDM. |
