## Supplemental File 3, and will be used for the link to the file on the preprint slide for "A realist review of medication optimisation of community dwelling service users with serious mental illness"

### Supplementary File 3: MEDIATE CMOC Coding Framework

| Overarching  Conceptual Theme | Parent Node | Child Node | Notes for researchers |
| --- | --- | --- | --- |
| Decision  Making | Coercion: |  | Pressure to follow advice against own wishes/may include threat of Mental Health Act |
|  | Independent decision-making |  |  |
|  |  | By provider | often when the provider deems the client incapable of making rational decisions |
|  |  | By client | includes autonomy, control, empowerment over own decisions |
|  | Shared decision-making: |  | evidence and options are discussed between client and provider-client’s input is equally valued and considered by the provider |
|  | Decisional factors- |  | often manifest during conflict/dilemma, new decision or change in condition |
|  |  | Available information: | includes insufficient or sufficient info or access to info |
|  |  | Family, peer roles/accountabilities | may be positive or negative |
|  |  | Cultural and social influences |  |
|  |  | Stigma | stigma against SMI |
|  |  | Other contextual considerations, | such as time and space for consultations, team-based care/inter-disciplinary approaches |
|  |  | Client insight or personal awareness-user/capacity/unwell |  |
|  | Therapeutic alliance/encounter/relationship-relational factors |  | motivational interviewing, informed consent |
|  |  | Patient approaches and perceptions |  |
|  |  | Provider approaches and perceptions |  |
|  |  | Strategies/interventions: |  |
| Med  Management  Interventions | Positive experiences and outcomes |  |  |
|  | Negative experiences and outcomes |  |  |
|  | Orals (+/- experiences) |  | Types of medications |
|  | Injectables [LAIs] (+/- experiences) |  |  |
|  | Theories |  | E.g., Example: Common sense or self-regulation theory in Pinfold  Stages of SDM and their barriers in Grunwald & Thompson  Personal identity formation |
|  | Background Information |  | with conceptual/operational definitions of key terms and stats |
